# Bringing Proportional Recovery into Proportion: Bayesian Hierarchical Modelling of Post-Stroke Motor Performance

**DOI:** 10.1101/19009159

**Authors:** Anna K. Bonkhoff, Thomas Hope, Danilo Bzdok, Adrian G. Guggisberg, Rachel L. Hawe, Sean P. Dukelow, Anne K. Rehme, Gereon R. Fink, Christian Grefkes, Howard Bowman

## Abstract

Accurate predictions of motor performance after stroke are of cardinal importance for the patient, clinician, and health care system. More than ten years ago, the proportional recovery rule was introduced by promising just that: high-fidelity predictions of recovery following stroke based only on the initially lost motor performance, at least for a specific fraction of patients. However, emerging evidence suggests that this recovery rule is subject to various confounds and may apply less universally than assumed by many.

We systematically revisited stroke outcome predictions by casting the data in a less confounded form and employing more integrative and flexible hierarchical Bayesian models. We jointly analyzed *n*=385 post-stroke trajectories from six separate studies – the currently largest overall dataset of upper limb motor recovery. We addressed confounding ceiling effects by introducing a subset approach and ensured correct model estimation through synthetic data simulations. Finally, we used model comparisons to assess the underlying nature of recovery within our empirical recovery data.

The first model comparison, relying on the conventional fraction of patients called *fitters*, pointed to a combination of constant and proportional to lost function recovery. Proportional to lost here describes the original notion of proportionality, indicating greater recovery in case of a more pronounced initial deficit. This combination explained only 32% of the variance in recovery, which is in stark contrast to previous reports of >80%. When instead analyzing the complete spectrum of subjects, model comparison selected a composite of constant and proportional to spared function recovery, implying a more significant improvement in case of more preserved function. Explained variance was at 53%.

Therefore, our data suggest that motor recovery post-stroke may exhibit some characteristics of proportionality. However, the levels of explanatory value were substantially reduced compared to what has previously been reported. This finding motivates future research moving beyond solely behavior scores to explain stroke recovery and establish robust single-subject predictions.

## Introduction

The science of recovery post-stroke has moved from comprehensive yet mainly anecdotal descriptions of patients’ trajectories (Newman, 1972; G. Broeks, *et al*., 1999) to increasingly larger studies aiming to create robust prediction models for individual outcome. Initial impairment status crystallized as one of the most predictive features, providing the foundation of the proportional recovery rule (Prabhakaran *et al*., 2008). According to this rule, the majority of stroke patients, considered *fitters* to the rule, recover about 70% of the initially lost function^1^ within the first few months after the initial event. *Fitters* and *non-fitters* to this proportional (to lost function) recovery rule have been defined in several ways in previous studies. For example, by choosing a discrete initial cut-off score, clustering initial and follow-up scores, or utilizing measures of corticospinal tract integrity. The proportional recovery rule, developed initially for Fugl Meyer assessment scores of the upper limb (Kundert *et al*., 2019), has since been extended to various functional domains. Numerous studies on recovery post-stroke claim to confirm proportional (to lost function) recovery of the upper limb (Zarahn *et al*., 2011; Byblow *et al*., 2015), the lower limb (Smith *et al*., 2017), language (Marchi *et al*., 2017), and neglect (Winters *et al*., 2017). Collectively, these studies consistently report high values of explained variance in cumulatively more than 500 participants, even as high as 94% (Winters *et al*., 2015).

Very recently, doubt has been placed on these effect size estimates (Hawe *et al*., 2019a; Hope *et al*., 2019). The concerns relate to the problem of mathematical coupling, when correlating an initial score and amount of change (Lord, 1956; Hayes, 1988; Chiolero *et al*., 2013). In case of this correlation, confounding effects arise due to one variable, e.g., change, directly or indirectly carrying information on the other one, e.g., the initial score. However, our concerns go beyond this basic observation by identifying a particularly severe coupling confound. This coupling confound may occur when the second (end) time-point is considerably less variable than the initial time point, leading to a small variability ratio. Such marked reductions of variability can result from ceiling effects at the second time-point, as frequently observed in stroke recovery data, based on the Fugl Meyer assessment (Gladstone *et al*., 2002).

Importantly, the severity of confounds due to small variability ratios is considerable (**Figure 1)**. As illustrated, the *degenerate regime* implies that the make-up of *Initial* and *End*, i.e., the numbers that comprise them, are irrelevant, and indeed, the relationship between *Initial* and *End* is irrelevant. Thus, testing proportional (to lost function) recovery when the variability ratio is very small is tautological – overwhelming evidence for it will, without question, be found. Because it is central to our argument, we name the degeneracy induced by a small variability ratio and the impact of concentration of data towards the ceiling *Compression enhanced Coupling*.

**Figure 1.**
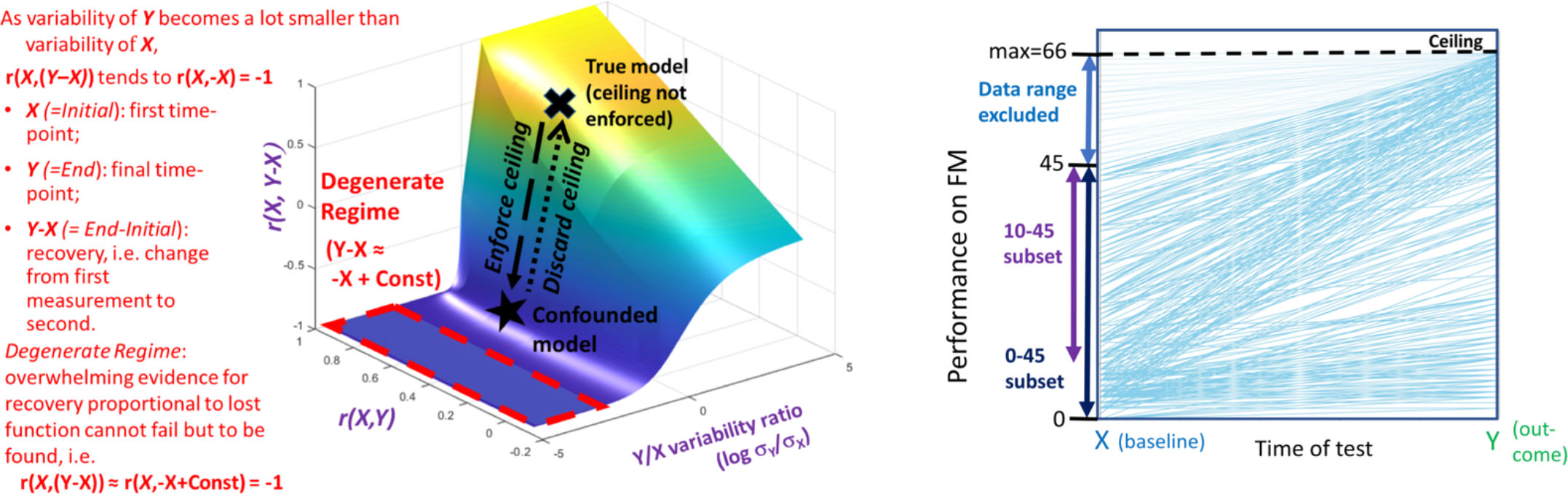
The confounded nature of *proportional to lost function* recovery assessments when the variability ratio is small and the resolution of confounds with ground truth simulations. Left (red text): The key correlation used to quantify *proportional to lost function* recovery, *r(X,(Y-X))*, effectively reduces to correlating *X* with *–X* when the variability at time-point one is substantially greater than at time-point two. **Middle (surface plot):** Full characterization of change correlations. Critically, the *degenerate regime* (region in red dashes) is almost entirely flat. Thus, when the variability of *Y* is substantially less than the variability of *X*, the correlation between *X* and *Y-X* degenerates to being very close to -1 in all cases. As a result, overwhelming, but spurious, evidence for recovery *proportional to lost function* cannot fail but to be found.

Once having identified this *Compression enhanced Coupling* confound, it is essential to consider whether it can be circumvented to enable an accurate assessment of the proportional recovery question. The latter is what we aimed to do in this article. Our logic was as follows.

1. The nature of recovery post-stroke cannot be meaningfully evaluated when there is a substantial ceiling effect at the second time-point (causing a small variability ratio).
2. By reducing data at ceiling, without incurring any new confounds, we can increase the variability ratio and lift the data set out of the *degenerate regime*, c.f. black annotations in **Figure 1**.
3. In a final step, we may then fit various de-confounded recovery models and perform model comparisons to assess the underlying mechanisms of stroke recovery.

The logic of synthetic data simulations is also shown (black text, arrows, and symbols). The black cross marks data generated from a ground truth model with known properties and initially without ceiling enforced. The position the cross marks has *r(X,Y-X)* and *r(X,Y)* as strong positive correlations and similar variabilities for *X* and *Y* (i.e., variability ratio around one, its log around zero). This, then, is synthetic data consistent with a *proportional to spared pattern* of recovery, similar, for example, to **panel [B] of Figure 2**. We then simulate the effect of ceiling, thereby, reducing the variability ratio, which, in turn, causes the data to exhibit a *proportional to lost pattern*, i.e., *r(X,Y-X)* becomes negative, and it moves in the surface plot (see black dashed arrow) towards the degenerate region; see black star. In particular, we will be able to show that by discarding data, we can return the synthetic data, via dotted black arrow, to ground truth position, i.e., black cross. Figure modified from (Hope *et al*., 2019). **Right: Depiction of subset approach**. All data sets are collapsed together for visualization purposes and presented as change across the two time-points (as per bottom row of Figure 2). We consider two subsets in this paper: *FM-initial* 10-45 (*Fitters* only) and *FM-initial* 0-45 (*Fitters & Non-fitters*). In both cases, the range from *FM-initial* 45-66 is excluded, in order to remove the ceiling. Data in this upper range is, therefore, shown in lighter colour.

**Figure 2.**
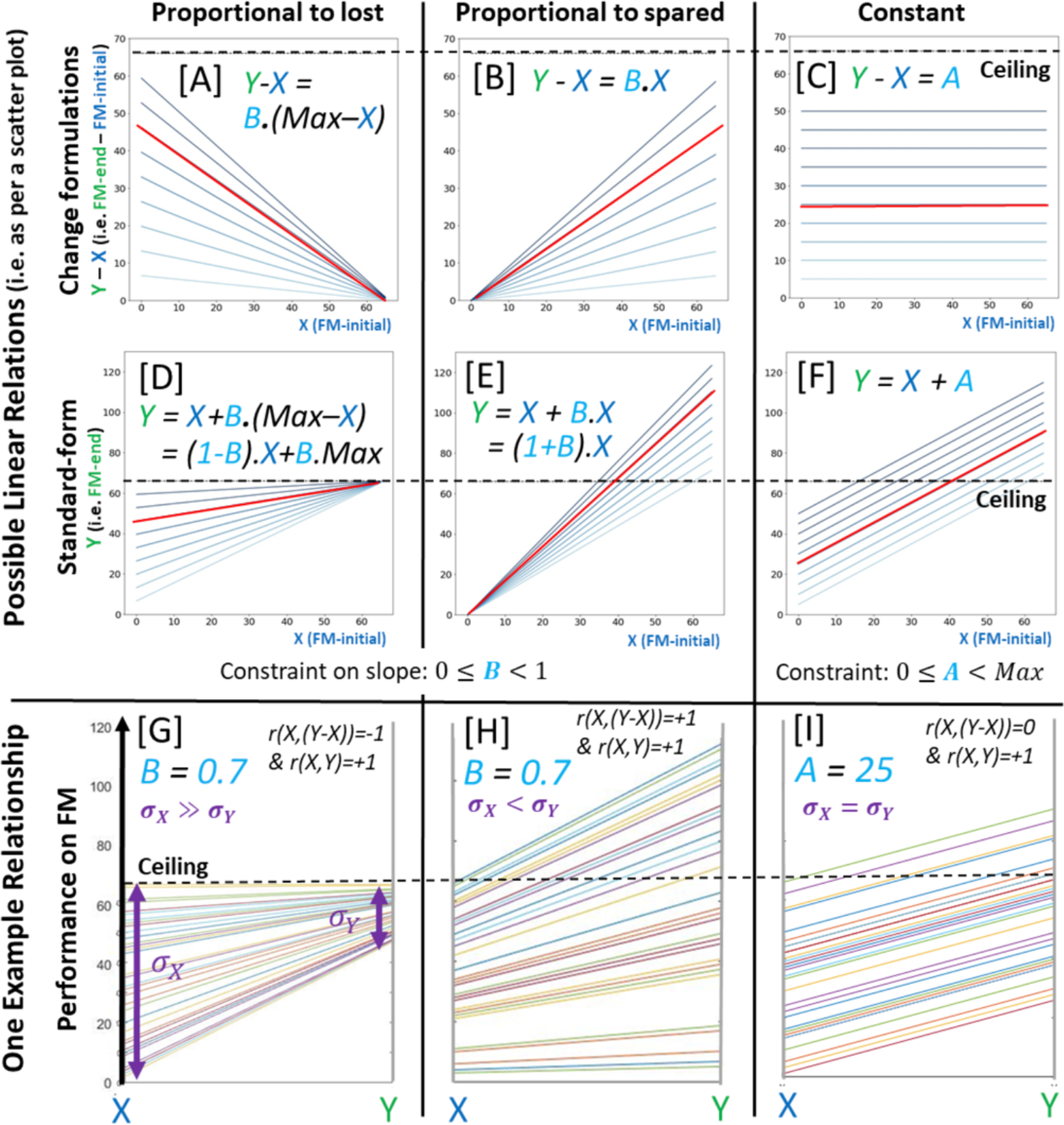
Various representations of the three forms of recovery we explore: *Proportional to lost function* (left column), *proportional to spared function* (middle column), and *constant* recovery (right column). Performance is inspired by the Fugl Meyer (FM) assessment of the upper limb, where 66 is the maximum value, providing a ceiling. However, the depictions here show the “true” underlying recovery pattern that one would obtain if there was no ceiling (thus, we extended the scales beyond the maximum). Top row panels (A, B & C) depict *proportional to lost, proportional to spared function*, and *constant* recovery to varying amounts (i.e., 10% proportional recovery, 20% proportional recovery etc.) under the typical change formulations. The horizontal axis presents initial FM scores, i.e., *X*, while the vertical axis stands for the change, i.e., *Y-X*. Middle row panels (D, E & F) depict the same linear relationships, but re-expressed as classical standard-form regressions, by merely moving the *X* variable to the right-hand side of the equation, and then, rearranging. In contrast to the top row, the vertical axis here represents the raw end score *Y*. Either formulation can be fit to the same data, resulting in corresponding intercepts and slopes, i.e., they can be transformed from one formulation to the other according to the relationships in supplementary materials section 1. Notably, the resulting variances explained would not typically be the same, since only the change formulation exhibits mathematical coupling; for a relevant discussion, see section Equality of Variances in Supplementary Appendix A of (Hope *et al*., 2019). Bottom row panels (G, H & I) depict one pattern of data that could arise from each of the recovery patterns. Each line in each plot indicates a single patient’s recovery trajectory, from the initial score (*X*), left side, to end score (*Y*), right side of the plot. For each pattern of recovery, the slope or intercept used to generate the data is shown; no noise term was included. These slopes or intercept reflect the linear relationship shown as a red line in the corresponding two panels above. The key correlations, *r(X,Y-X)* and *r(X,Y)*, that hold are also shown, along with the relationship between standard deviations of *X* and *Y*.

In the present study, we revisited data at ceiling by creating subsets, i.e., we excluded a (varying) range of stroke participants with the highest scores at the *Initial* time-point. This procedure also reduced the ceiling effect at the second *End* time-point (**Figure 1)**. Critically, we validated this subset procedure in synthetic data experiments (Gelman and Hill, 2006). That is, we generated data with known ground truth and simulated three candidate explanations of recovery post-stroke (**Figure 2**): a) *proportional to lost function*; b) *proportional to spared function*; and c) *constant* recovery. *Proportional to lost function* is the familiar pattern, where a more profound initial deficit implies a more significant recovery. *Proportional to spared function* is the opposite pattern and encapsulates the notion that the more preserved the function at time-point one, the greater the potential for an individual to recover. *Constant* recovery formalizes the idea that initial severity has no impact on recovery, which is the same size, whatever the initial functional deficit.

Subsequently, we aimed to assess recovery patterns in empirical stroke data. Therefore, we aggregated a substantial body of data, i.e., 385 individual post-stroke recoveries, across a range of representative studies focused on upper limb deficits measured as Fugl Meyer scores (Buch *et al*., 2016; Byblow *et al*., 2015; Feng *et al*., 2015; Guggisberg *et al*., 2017; Winters *et al*., 2015; Zarahn *et al*., 2011). We employed state-of-the-art hierarchical Bayesian models, enabling us to incorporate data from these various sources while accounting for inter-study variability and explicitly modeling the full probability landscape. Crucially, these models also permitted conducting overall model comparisons. We considered the three change models mentioned before and shown in **Figure 2** – *proportional to lost, proportional to spared*, and *constant recovery*. We also included classical standard-form linear regression, the most general of the models, which determined whether linear relationships outside our three candidate models explained the data any better. Lastly, we evaluated how well each of these models explained inter-individual differences in recovery.

The core objective of this paper was to respond to the confounded nature of assessments of behavioral recovery from stroke, particularly from upper-limb deficits. We did so by discarding data to unconfound the fitting problem. We then invoked Bayesian models to answer the substantive scientific question: what biological mechanisms best explain the data on recovery of upper-limb performance after stroke and with what effect size?

## Material & Methods

### Participants and clinical data

The analyses of post-stroke upper limb function were based on a sample of 385 acute stroke participants originating from six different studies on stroke recovery (Buch *et al*., 2016; Byblow *et al*., 2015; Feng *et al*., 2015; Guggisberg *et al*., 2017; Zarahn *et al*. 2011). Details on data acquisition are given in the supplementary material. In brief, we used anonymized data available from Zarahn *et al*. (2011) and Guggisberg *et al*. (2017) and combined it with secondary data from (Hawe *et al*., 2019*a*). Therefore, we had individual-level information on Fugl Meyer (FM) scores assessing upper limb motor performance in the acute as well as chronic stage (three to six months after the event; Nakayama *et al*., 1994). A minimum score of 0 implies no preserved and 66 maximal motor performance (Fugl-Meyer *et al*., 1975). In line with previous research (Feng *et al*., 2015), we split the stroke subject samples into *fitters* and *non-fitters* to the classic proportional recovery rule (Prabhakaran *et al*., 2008) based on their initial scores (Non-Fitters: *FM-Initial*≤10 points, Fitters: *FM-Initial*>10 points). The first set of analyses were focused exclusively on *fitters*, and subsequent analyses highlighted findings on the entire sample, i.e., *fitters* and *non-fitters*. As all of the data has been published previously, ethics approvals had been granted for all individual primary studies.

### Bayesian hierarchical modelling of motor stroke outcome

A hierarchical Bayesian framework was employed to allow for the balanced incorporation of data from various sources and facilitate model comparisons. More precisely, we built Bayesian multilevel (hierarchical) linear regression models with varying intercepts and slopes (Gelman, 2006). Therefore, each of the six considered studies was characterized by estimated full probability distributions of intercept and slope parameters – rather than simple best-fit, maximum likelihood parameter estimates as usually employed in recovery studies. Retaining study-specific information in this statistical way was an essential step to address potential differences between studies. These differences are conceivable due to independent data collection, involving study sites in different countries, and likely minor variations in therapy regimens. Nonetheless, given that each of the included studies considered similar measures at similar time points from broadly similar participants, information was also pooled across the various studies and a set of hyperparameters, i.e., across-study intercept and slope, was derived (Bzdok *et al*., 2019). Thus, intercepts and slopes had two levels that carefully captured across-study versus individual variation in the eligible stroke studies.

The outcome variables that we sought to predict were either the raw *FM-end* score or *Change* (i.e., *FM-end – FM-initial*). We thus created a likelihood function for the outcome and linked it to the priors of our predictor variables, i.e., either the unaltered *FM-initial* score or *Potential* (*FM-maximum – FM-initial*), through one of five different models:

**The (classical) standard-form regression model:** *FM-end = b*.*FM-initial + a* (with intercept *a* and slope *b*)

**The change model:** *Change = Potential*.*B + A*, with *Change = FM-end – FM-initial* and *Potential = FM-maximum – FM-initial*, with FM-maximum = 66.

The change model was more precisely framed in three different ways expressing various conceivable recovery models, which we highlighted in the Introduction (see panels [A,B,C] of **Figure 2**): The **classical proportional to lost function recovery model:** *Change = Potential*.*B*, a **proportional to spared function recovery model:** *Change = FM-initial*.*B* (which takes the raw initial score), and a **constant recovery model:** *Change = A*. Once fitted, we determined whether the obtained models were truly proportional to lost, spared, or constant recovery by assessing the parameter settings, where 0 ≤ *B*<1 for the proportional to lost or spared function recovery and where 0 ≤*A*< *Max*∧ *Max*= 66for the constant recovery model.

A critical step in any Bayesian analysis is the specification of prior beliefs. We attenuated the effects of priors and simultaneously increased the influence of the actual data by choosing simple, weakly informative Gaussian priors (i.e., with large standard deviations) for all of the (hyper-)parameters and half-Cauchy priors for corresponding variance terms.

The full Bayesian model is specified below: Dependencies between variables are indicated with arrows, observed variables are in grey boxes, and the distributional definition of variables are shown on the right. *M* denotes the number of studies analyzed and *N(i)* the number of subjects in each study.

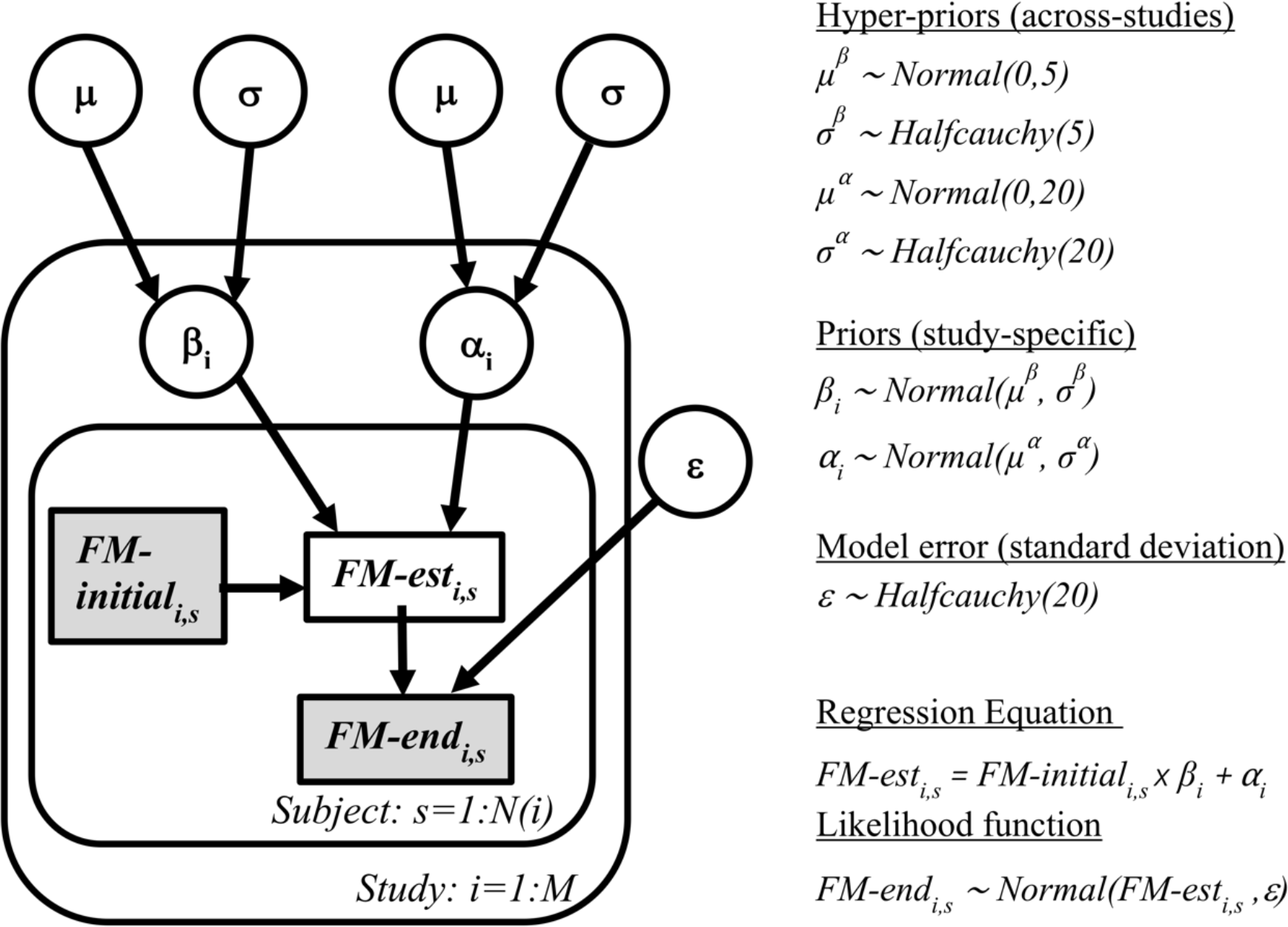

Inference: The analytical derivation of posterior distributions is either computationally very expensive and challenging or not possible, as it requires the integration over thousands of unknown parameters. We thus deployed a recent Monte Carlo Markov Chain algorithm, the No U-Turn Sampler (NUTS), that does not compute the posterior distribution directly, yet draws samples from it in a stochastic way (Hoffman & Gelman, 2014, setting: draws=2000, n_init=1000; for quality assurance and to check for convergence: initially 4 chains, then 1 chain for final analyses). Marginal posteriors are given as mean and 95%-credible intervals. Posterior predictive checks were run to analyze the model performance, i.e., we predicted *FM-end* or *Change* scores based on parameter drawings from the posterior. In this way, we could assess whether data originating from our fitted hierarchical model resembled data from the true underlying distribution. We compare predicted means to the actual sample means and finally compute R-squared values as a measure of explained variance.

### Synthetic data simulation experiments

Before performing Bayesian model comparisons, we first conducted data simulations (Gelman & Hill, 2006, Chapter 8, p. 155) and synthetically generated data based on ground truth model. This procedure enabled us to better understand and validate statistical procedures. Thus, these simulations enabled us to test strategies to ensure correct model estimation despite the effects of noise and ceiling. The ceiling effect is most similar to having right-censored data, i.e., just knowing that individual data points are at a maximum value, which they, however, may exceed by an unknown amount.

We proceeded in the following way: We selected *constant, proportional to lost*, and *proportional to spared function* recovery as “true” models. These models were re-arranged to obtain the standard-form classical regression, directly linking X and Y, and enabling us to generate Y-values from those of X (c.f. **supplementary materials section 1** for details and corresponding proofs of these transformations). To consider different degrees of recovery, we assessed these in 10% steps from 10–90% of the proportional recoveries and in steps of 5 from 5–50 points for constant recovery. We then entered the empirical *FM-initial* scores of all *fitters* (n=243) in one of the “true” models, added noise, and enforced ceiling (for details on these procedures, c.f. **supplementary materials section 4**). The final and critical step then was to fit a new linear regression model to the simulated data and compare the estimated parameters for intercept and slope to the given “true” parameters to answer whether it was still possible to estimate the “true” model after alterations by noise and ceiling. Therefore, we assessed the probability that the “correct” (ground truth) parameter values fell in the confidence intervals of the parameter values inferred by the fitting process and evaluated the percentage meeting 68% and 95% confidence intervals (Gelman & Hill, 2006). Also, we tracked the ratio of the standard deviations *FM-end/FM-initial*, Pearson correlations of *FM-initial* & *FM-end*, as well as *FM-initial* & *Change* and the number of simulated subjects at absolute ceiling, i.e., at an FM of 66 (maximum score). We are referring to “absolute” ceiling as further subjects might present with confounded scores that are compressed toward ceiling, yet not at *FM-end*=66.

Aiming to reduce ceiling and thus its confounding effect, we implemented a subset approach by limiting the data simulations to specific *FM-initial* ranges, i.e., subset 1) *FM-initial* 10–60, 2) *FM-initial* 10–50, 3) *FM-initial* 10–45, and 4) *FM-initial* 10–40, and evaluated its effect on subsequent model estimation. This enabled us to assess in synthetic data, which subset approach gave us the best trade-off between retrieval of correct model and parameter settings, and size of the remaining data. In each of the described scenarios, simulations were repeated 1000 times.

### Final model comparisons

#### Fitters only

We initially focused on the *fitters* (*FM-initial>10*) portion of the data. Relying on the simulation results, we constructed Bayesian hierarchical models for the standard-form regression, the *proportional to lost function*, and *proportional to spared function* as well as the *constant* recovery models in the subset *FM-initial* 10–45 and conducted a Bayesian model comparison. We focused on this subset, as it represented an optimal compromise between mitigating ceiling effects and retaining as many subjects in the analysis as possible. Results for the subsets *FM-initial* 10–40 and *FM-initial* 10–50 are provided in supplementary materials.

Despite our dataset being as voluminous as currently possible, a potential limitation is that, for some of the studies included, we relied upon values extracted from published figures. This process missed 68 subjects because multiple points sat on top of one another in scatter plots. To account for these missing values and determine an upper bound of the R-squared for the winning model in our model comparison, we repeatedly (1000 times) took 68 random draws from the available *FM-initial* distribution. We discarded values not in the range 10–45 and placed the remaining values on the predicted linear fit (i.e., assuming perfect prediction by the standard-form model). By these means, we obtained an average R-squared value, which can be considered an *upper bound* when correcting for missing values.

#### Fitters and non-fitters

In the final analyses, we jointly investigated data on *fitters* (*FM-initial*>10, n=243) and *non-fitters* (*FM-initial*≤10, n=142) by fitting the four competing models outlined before. Once again, we ran analyses with a decreased upper limit for *FM-initial* scores to prevent confounding by ceiling (*FM-initial* cut-off: 45).

### Statistical analyses

The main analyses were conducted in a Bayesian hierarchical framework. The central inferential question is a model comparison. Specifically, we determined the models best describing the data based on their Leave-One-Out-Cross-Validation (LOOCV) (Vehtari *et al*., 2017), with LOOCV being a critical out-of-sample test of model fit – indicating whether our findings are likely to hold up in future studies. Model comparisons based on the widely applicable information criterion (WAIC) are also given in supplementary materials. The WAIC represents a principled means of weighing goodness-of-fit against model complexity (i.e., number of effective parameters) (Watanabe, 2013). Additionally, we report R-squared values, the standard measure of the effectiveness of models at explaining within-sample variability.

### Data availability

The recovery data as well as jupyter notebooks (python 3.7, primarily software package pymc3, (Salvatier *et al*., 2016)) employed in this study are available from the authors on reasonable request.

## Results

### Descriptive statistics

Individual studies, as well as the joint distributions of *Initial* and *End* FM scores, median values, and quartiles, are illustrated in **Figure 3**. Further characteristics, such as size, mean age, and sex of each study are summarized in **Table 1**.

**Table 1.**
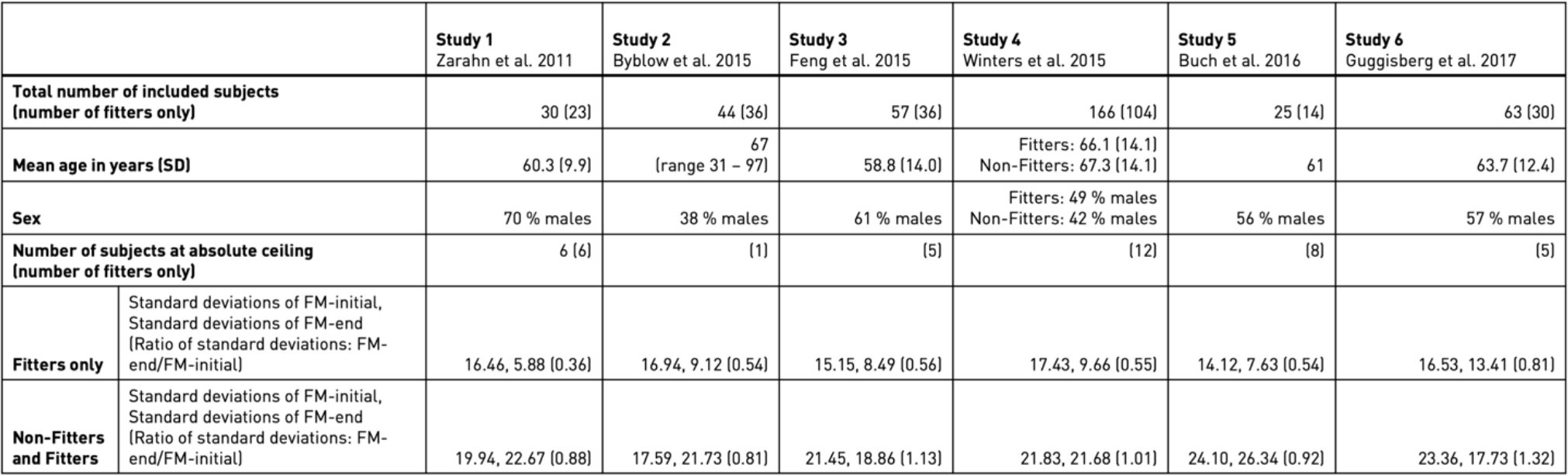
Descriptive statistics of each of the six studies included.

|  |  | Study 1<br>Zarahn et al. 2011 | Study 2<br>Byblow et al. 2015 | Study 3<br>Feng et al. 2015 | Study 4<br>Winters et al. 2015 | Study 5<br>Buch et al. 2016 | Study 6<br>Guggisberg et al. 2017 |
| --- | --- | --- | --- | --- | --- | --- | --- |
| Total number of included subjects<br>(number of fitters only) |  | 30 [23] | 44 [36] | 57 [36] | 166 [104] | 25 [14] | 63 [30] |
| Mean age in years (SD) |  | 60.3 [9.9] | 67<br>[range 31 – 97] | 58.8 [14.0] | Fitters: 66.1 [14.1]<br>Non-Fitters: 67.3 [14.1] | 61 | 63.7 [12.4] |
| Sex |  | 70 % males | 38 % males | 61 % males | Fitters: 49 % males<br>Non-Fitters: 42 % males | 56 % males | 57 % males |
| Number of subjects at absolute ceiling<br>(number of fitters only) |  | 6 [6] | [1] | [5] | [12] | [8] | [5] |
| Fitters only | Standard deviations of FM-initial,<br>Standard deviations of FM-end<br>(Ratio of standard deviations: FM-end/FM-initial) | 16.46, 5.88 [0.36] | 16.94, 9.12 [0.54] | 15.15, 8.49 [0.56] | 17.43, 9.66 [0.55] | 14.12, 7.63 [0.54] | 16.53, 13.41 [0.81] |
| Non-Fitters<br>and Fitters | Standard deviations of FM-initial,<br>Standard deviations of FM-end<br>(Ratio of standard deviations: FM-end/FM-initial) | 19.94, 22.67 [0.88] | 17.59, 21.73 [0.81] | 21.45, 18.86 [1.13] | 21.83, 21.68 [1.01] | 24.10, 26.34 [0.92] | 23.36, 17.73 [1.32] |

**Figure 3.**
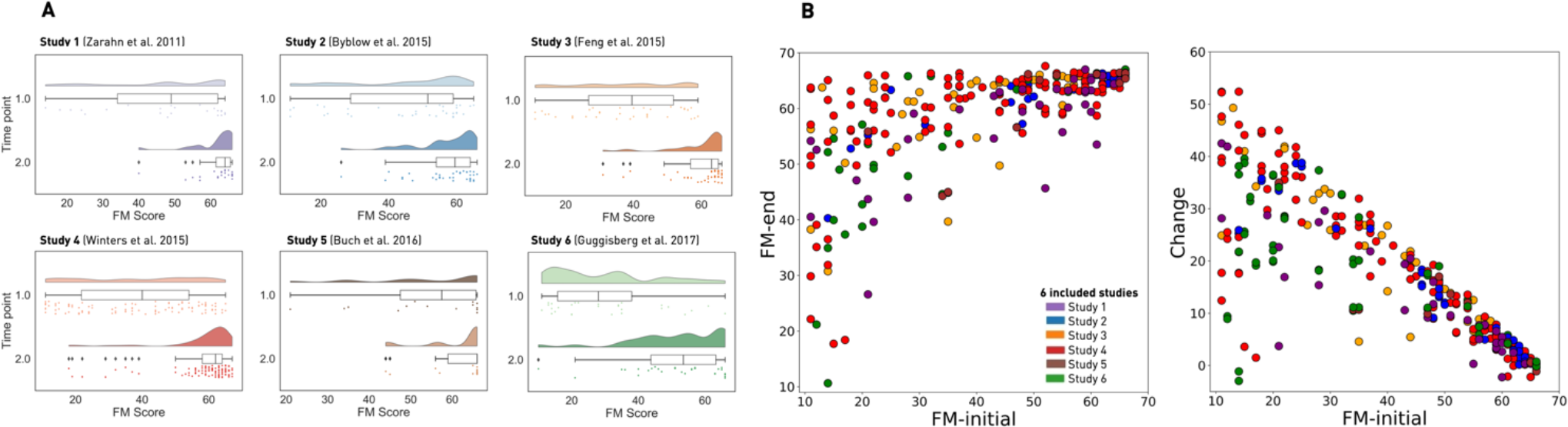
Initial and follow-up upper limb motor performance, individually for each of the six studies included and aggregated across studies. A. Raincloud plots of the initial (time point 1) and follow-up (time point 2) Fugl Meyer (FM) assessment scores. These measured the upper limb impairment post-stroke of all *fitters*, i.e., those with *FM-initial* > 10. Each of the six studies included is displayed separately and uniquely color-coded. The upper row within each study’s plot visualizes the distribution of scores. The second row summarizes the same data in a boxplot (i.e., median, upper and lower quartiles, whiskers extending to the entire range of data, outliers indicated as separate dots). Lastly, the third row displays raw individual data points. While initial FM score distributions are more homogeneous across the entire range (i.e., more uniform), distributions at the second time point are narrower and – to varying degrees – more pronounced at the upper end of the FM assessment scale, i.e., skewed. Of note, Feng *et al*. (2015) only considered stroke subjects with initial FM scores lower than 60. Guggisberg *et al*. (2017) included more subjects with lower initial scores and scheduled the second assessment sooner on average, which likely underlies the more widespread and less skewed distribution of *FM-end* scores. The code for raincloud plots relies on (Allen *et al*., 2019). **B. Entirety of aggregated individual FM assessment scores of all stroke subjects defined as *fitters* (*FM-initial* > 10)**. Scores are jittered on the vertical axis for visualization only. Left: *FM-initial* vs. *FM-end*, clearly depicting the increased density for follow-up scores close to and at ceiling, i.e., *FM-end* = 66. Right: *FM-initial* vs *Change* (*FM-end – FM-initial*). Included studies are color-coded as before, c.f. legend.

### Bayesian posterior distributions for stroke recovery prediction

The hierarchical standard-form regression and change models built on the entirety of *fitters* could be well estimated, as indicated by the convergence of four independently sampled (Monte Carlo Markov) chains for the posterior estimates of model parameters. Additionally, the predicted posterior mean was evenly distributed around the actual sample mean, indicating that the model can reproduce patterns occurring in the real data (c.f. **Supplementary Figure 2**). **Figure 4** illustrates the marginal posteriors for the across-study and study-specific intercept and slope parameters, arising from one final chain. Furthermore, **Figure 5** highlights the joint posterior densities for intercept and slope parameters. A striking finding for both models was the dispersion of the individual studies’ intercepts and slopes: For the change model, the across-study mean for the slope was 0.64, thus specifying a proportional recovery of 64% for all six studies combined. However, the individual posterior means for slopes fell in the range between 54% (Guggisberg *et al*., 2017) and 70% (Feng *et al*., 2015). The six slopes also followed two general patterns, with three studies featuring lower and three studies higher proportional recovery amounts. As expected, the explained variance of the change model markedly surpassed that of the standard-form regression model (Predictive posterior check (PPC): R-squared: 70.8% vs. 42.7%), demonstrating the problematic inflation due to mathematical coupling highlighted in Hawe *et al*. (2019a) and Hope *et al*. (2019). Also, this dispersion of effect sizes coincided with a small ratio of standard deviations *FM-end/FM-initial*, totaling 0.57, which, at least partially, resulted from the number of subjects reaching absolute ceiling at follow-up: 37 (15.2%). In sum, these are the canonical properties of *Compression Enhanced Coupling*.

**Figure 4.**
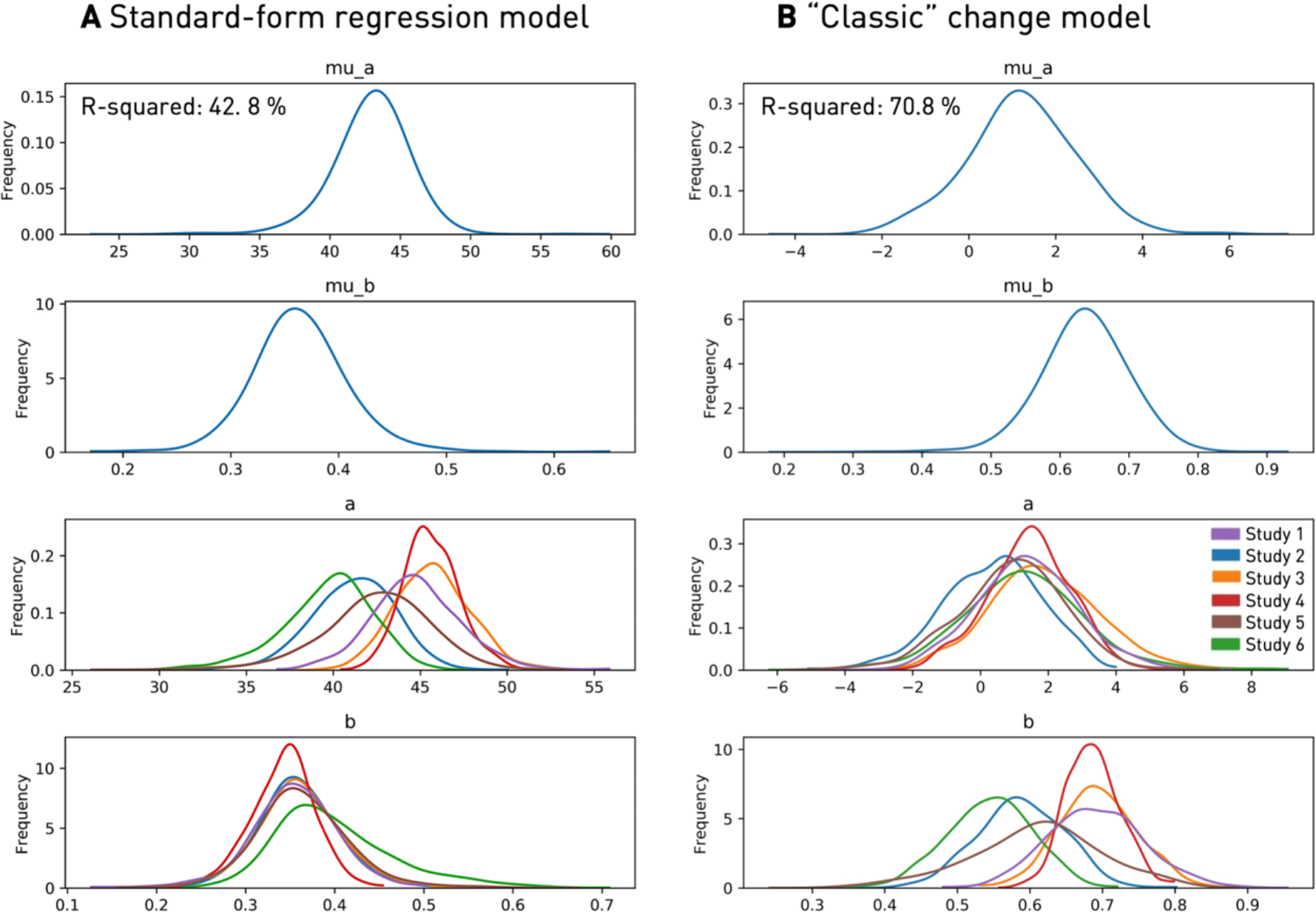
Bayesian hierarchical model: Marginal posteriors of intercepts and slopes. **A. Standard-form regression model.** Follow-up FM scores are estimated according to *FM-end* = b.*FM-initial* + a. *Across-study* intercept (“mu_a”) and slope (“mu_b”) are depicted in the upper two rows. The bottom two rows visualize the lower level of the hierarchy: Varying intercepts (“a”) and slopes (“b”), individually per included study (c.f. legend for study-specific color coding). **B. “Classic” *proportional to lost function* change model**. The outcome measure of interest here is the change between the initial and end FM scores, estimated based on *Change* = B.*Potential* + A with *Change = FM-end – FM-initial* and *Potential* = 66 – *FM-initial*. Marginal posteriors of individual slopes show a marked variability, ranging from 54% to 70% of proportional recovery. This might indicate a generally high inter-study variability. The inflation of explained variance due to mathematical coupling and *Ceiling enhanced Coupling* is demonstrated by an R-squared value of 70.8% for the change model that exceeds the one from the standard-form regression model by 28%. Please note that unlike our presentations of *proportional to lost function* elsewhere, we include an intercept here to maximize fit, although, as can be seen, fitting generates an intercept very close to zero.

**Figure 5.**
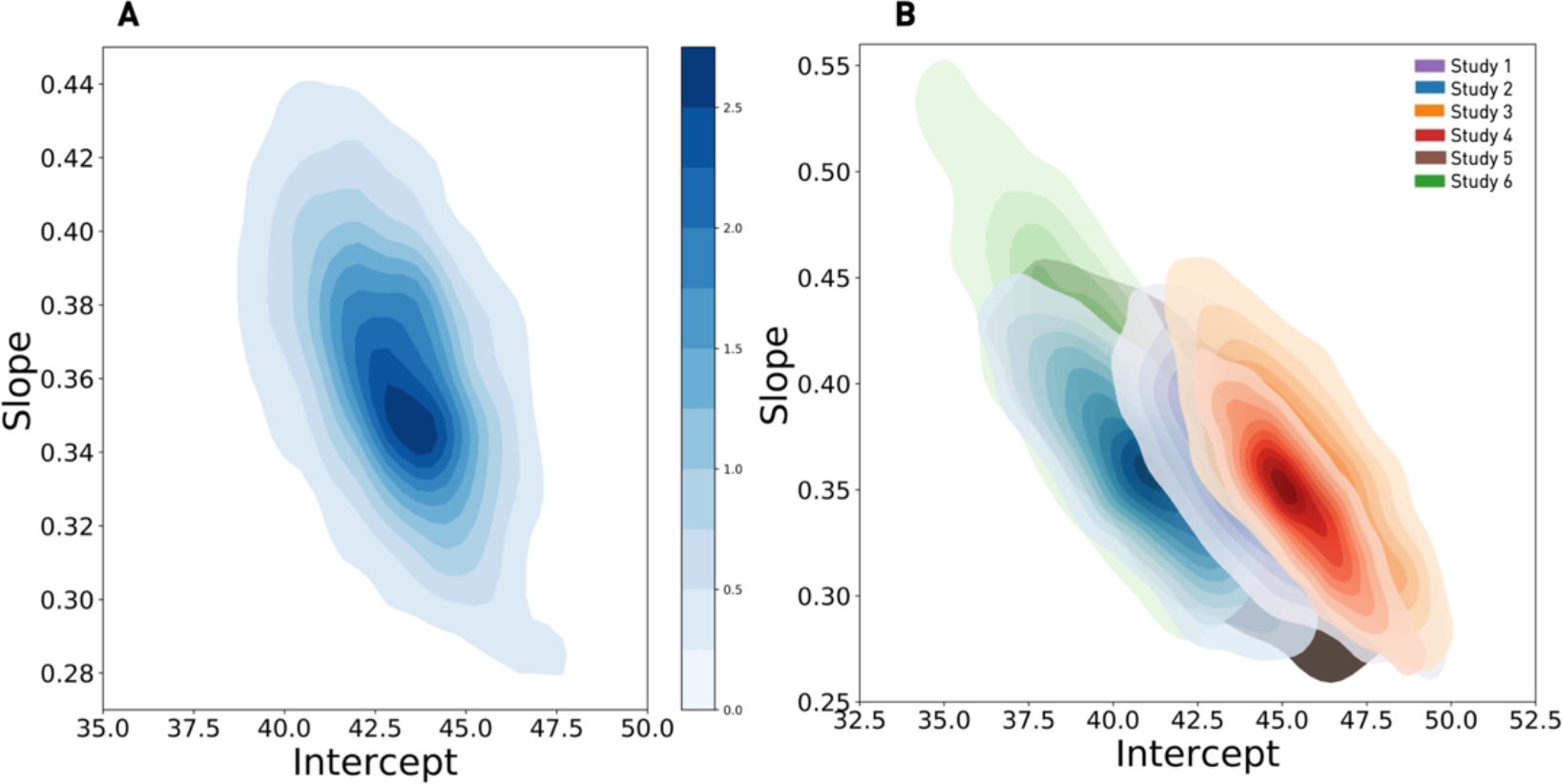
Model estimated within- and between-study differences in motor recovery based on the standard-form regression model. **A. Aggregated motor studies:** We here display the joint distribution between the *across-study* intercept – which can be described as the average motor outcome for an *FM-initial* score of zero – and the *across-study* slope – equivalently framed as performance gain dependent on *FM-initial*. Therefore, the plot illustrates the joint posterior densities for the included hyperparameters, with the marginal posterior for the intercept ranging from 37.0 to 47.4 and from 0.28 to 0.44 for slopes (95% credibility intervals). **B. Individual motor studies:** The figure pictures the joint density for combinations of intercepts and slopes that are plausible, given the visited data of the six included studies. It particularly highlights the relationship between sample size and width of credibility intervals, as larger studies present with narrower intervals. C.f. legend for study-specific color-coding.

### Synthetic data simulation experiments

Leveraging synthetic data simulations in the sample of *fitters* (Gelman and Hill, 2006) facilitated the detailed study of confounding effects of noise and ceiling as well as hypothetical conclusions when assuming three conceivable ground truth models: *proportional to lost function, proportional to spared function*, as well as *constant* recovery.

Detailed descriptions, as well as tables of the data simulations, are given in supplementary materials section 7. In sum, the inclusion of noise did not impede the correct model and parameter estimation, as indicated by met confidence intervals; that is >68% and >95% of the estimated parameters were in the 68%- and 95%-confidence intervals of the true model. This situation changed markedly with the introduction of a ceiling: correct model and parameter estimation deteriorated in parallel to the increase of subjects at absolute ceiling and with growing amounts of recovery (e.g., going from 10% of proportional recovery to 20%). The highest numbers of subjects at ceiling (n=110) arose for a constant recovery of 50 points. We could observe the effects of *Compression enhanced Coupling* since tracked Pearson correlations of *FM-initial* & *Change* increasingly surpassed those of *FM-initial* & *FM-end* after enforcing ceiling.

Crucially, any of the conceivable ground truth models introduced here closely resembled a *proportional-to-lost-function* recovery model once a ceiling was enforced, thus mirroring the scenario encountered with the FM score which has a finite maximum score of 66 (Gladstone *et al*., 2002). Hence, when considering all *fitters* and disregarding any potential ceiling effects, there was only one possible conclusion: Data would follow a *proportional to lost function* recovery regardless of the real mechanism driving recovery (**Figure 6**).

**Figure 6.**
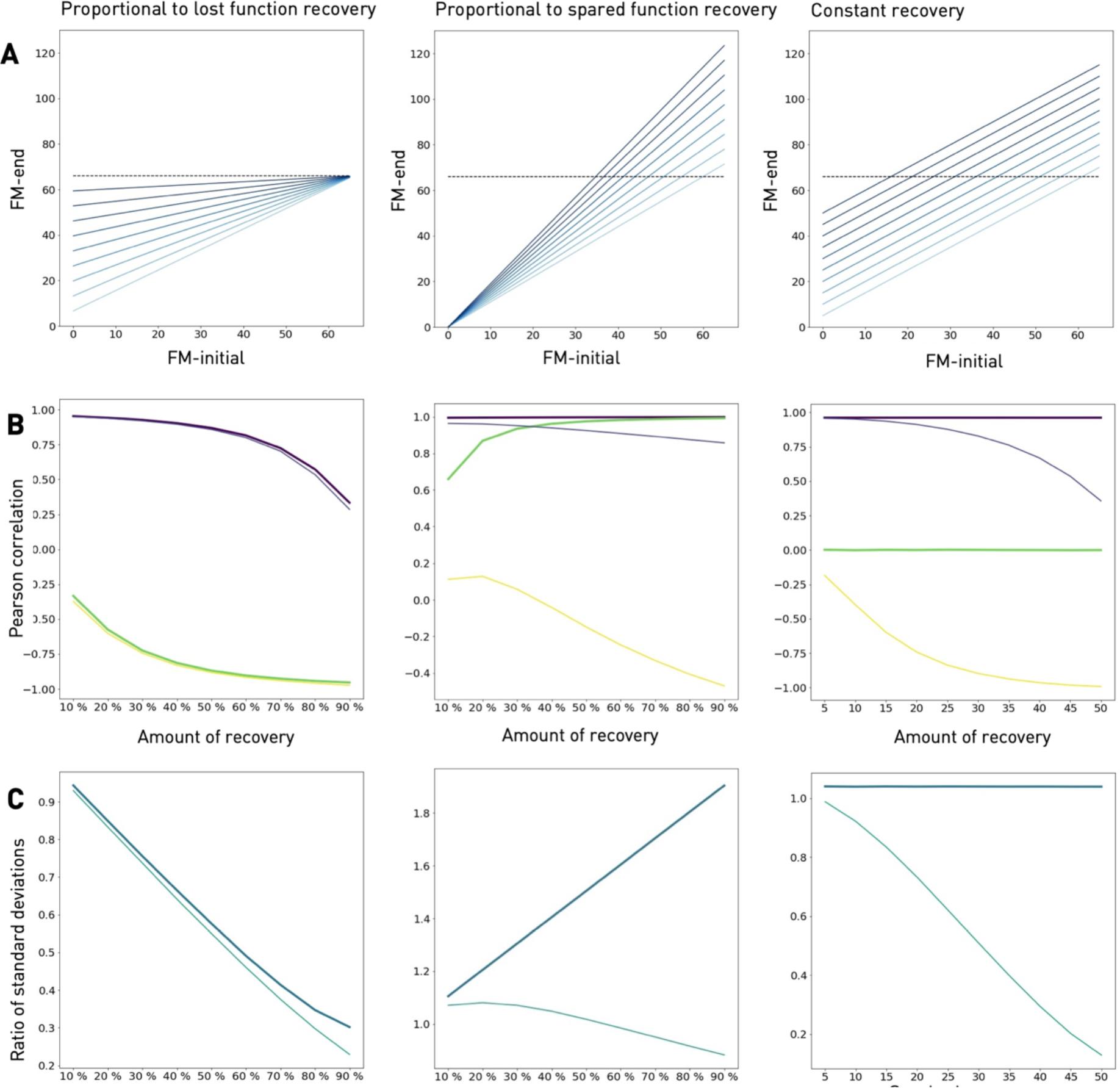
Synthetic data simulations of *proportional to lost function* (left column), *proportional to spared function* (middle column), and *constant* recovery (right column). A. Space of ground truth models. Illustrations of idealized recovery profiles (i.e., without random noise and any ceiling enforced) for standard-form formulations from 10% to 90% of proportional, and 5 to 50 points of constant recovery. The dashed lines indicate maximum Fugl Meyer scores of 66. While no subject would exceed the maximum in an idealized *proportional to lost function* scenario (note: this could be altered by noise), *proportional to spared function* and *constant* recovery naturally cause a substantial number of subjects at absolute ceiling (*FM-end* = 66). **B. Trajectories of Pearson correlation between *FM-initial (X) & FM-end* (Y) and *FM-initial (X) & Change (Y-X)***. Thick purple lines picture the course of *r(X,Y)* before and thin purple lines the course of *r(X,Y)* after enforcing ceiling for increasing amounts of recovery. Respective courses for *r(X, Y-X)* are depicted in green (before) and yellow (after ceiling). *Proportional to lost function recovery:* Starting at almost the maximum of one, the correlation between *FM-initial* & *FM-end* decreases the higher the amount of proportional recovery, while the correlation for *FM-initial* & *Change* becomes more negative and finally exceeds the one of *FM-initial* & *FM-end* in absolute terms, demonstrating the effect of mathematical coupling. Ceiling exhibits a minor amplification of this effect only. *Proportional to spared function recovery:* Without any ceiling, correlations of *FM-initial* & *FM-end*, as well as *FM-initial* & *Change*, are close to 1. The latter changes dramatically after enforcing ceiling: The correlations of *FM-initial* & *Change* are now decreasing monotonically, becoming negative in sign and reminiscent of *proportional to lost function. Constant recovery:* The correlation between *FM-initial* & *FM-end* is close to 1, while *FM-initial* and *Change* are not correlated. After ceiling is enforced, patterns closely resemble the ones for *proportional to lost function*: The correlation of *FM-initial* & *FM-end* decreases monotonically, yet stays positive, the one between *FM-initial* & *Change* decreases and becomes almost negative one for high levels of constant recovery. **C. Trajectories of ratios** σ(***FM-end)/***σ(***FM-initial)***. Thick lines in turquoise color mark trajectories before, thin lines in turquoise trajectories after enforcing ceiling. *Proportional to lost function recovery:* Ratios of standard deviations are decreasing from approx. 1 to 0.2, ceiling exhibits only minor effects. *Proportional to spared function recovery:* Trajectories differ markedly depending upon ceiling: Ratios are greater than one and increase before ceiling and decrease to values smaller than one after ceiling is enforced. *Constant recovery:* Once again, the presence of a ceiling substantially alters the trajectories of standard deviation ratios: While they remain close to 1 before enforcing ceiling, they show a steep decrease after enforcing ceiling. Importantly, whenever the variability ratio, expressed as *σ(FM-end)/σ(FM-initial)* in the bottom row C, decreases, the correlation between *FM-initial* & *Change* in the middle row B also decreases. This is consistent with *Compression enhanced Coupling*. It also implies that data enters the degenerate regime of figure 1, and any underlying model will “look like” *proportional to lost function* recovery.

However, estimation performance gradually improved again, when running the simulations in subsets of *fitters*, i.e., considering only those below a certain cut-off of initial scores. The most stringent subset of *FM-initial* 10–40 performed the best in terms of estimating the true intercepts and slopes for all of the conceived models. However, in order to choose an appropriate subset range for intended consecutive model comparisons, we tried to find the optimal balance of reducing possible confounds, while also retaining as many subjects in the analysis as possible. The subset *FM-initial* 10–50 was only capable of recovering *proportional-to-spared-function* up to a proportion of 30% and 15 points of *constant* recovery, which we did not judge to be sufficient. On the other hand, we could only keep three studies (92 subjects out of 385 in all the studies, i.e., 24%) in the case of the subset *FM-initial* 10–40, which seems an inefficient use of hard-won empirical data and reduction to too few patients. Therefore, we established a further subset *FM-initial* 10–45 to combine the advantages of both subsets, *FM-initial* 10–50 and 10–40. Based on the subset *FM-initial* 10–45, we were able to estimate the entire range of *proportional-to-lost-function*; up to 40% of *proportional to spared function*, and up to 20 points of *constant* recovery. Besides, we were able to recover the true “space” of the generating model for all proportional to spared function models, c.f. section 5 of supplementary material. This was done using recovery data from 118 subjects originating from four studies (out of 385, therefore 31%). Hence, we decided to focus upon the subset *FM-initial* 10–45 for model comparisons on the human data (results for *FM-initial* 10–50 and 10–40 are presented in supplementary materials).

### Final model comparisons (on human data)

#### *Fitters* in the subset of FM-initial 10 – 45

The studies by Zarahn *et al*. (2011) and Buch *et al*. (2016) were excluded from these analyses since they had fewer than ten subjects in the range of *FM-initial* 10–45, which would lead to a substantial deterioration in accuracy when sampling from the posterior. Relying on the remaining 118 subjects (out of 385, 31%; 6 subjects were at absolute ceiling, 5.1%; variability ratio was 1.12), we successfully sampled posteriors for the (unconstrained) standard-form regression model, and change-form versions *of proportional to lost function, proportional to spared function*, and *constant-recovery* models. Resulting distributions for the marginal posteriors are displayed in **Figure 7**.

**Figure 7.**
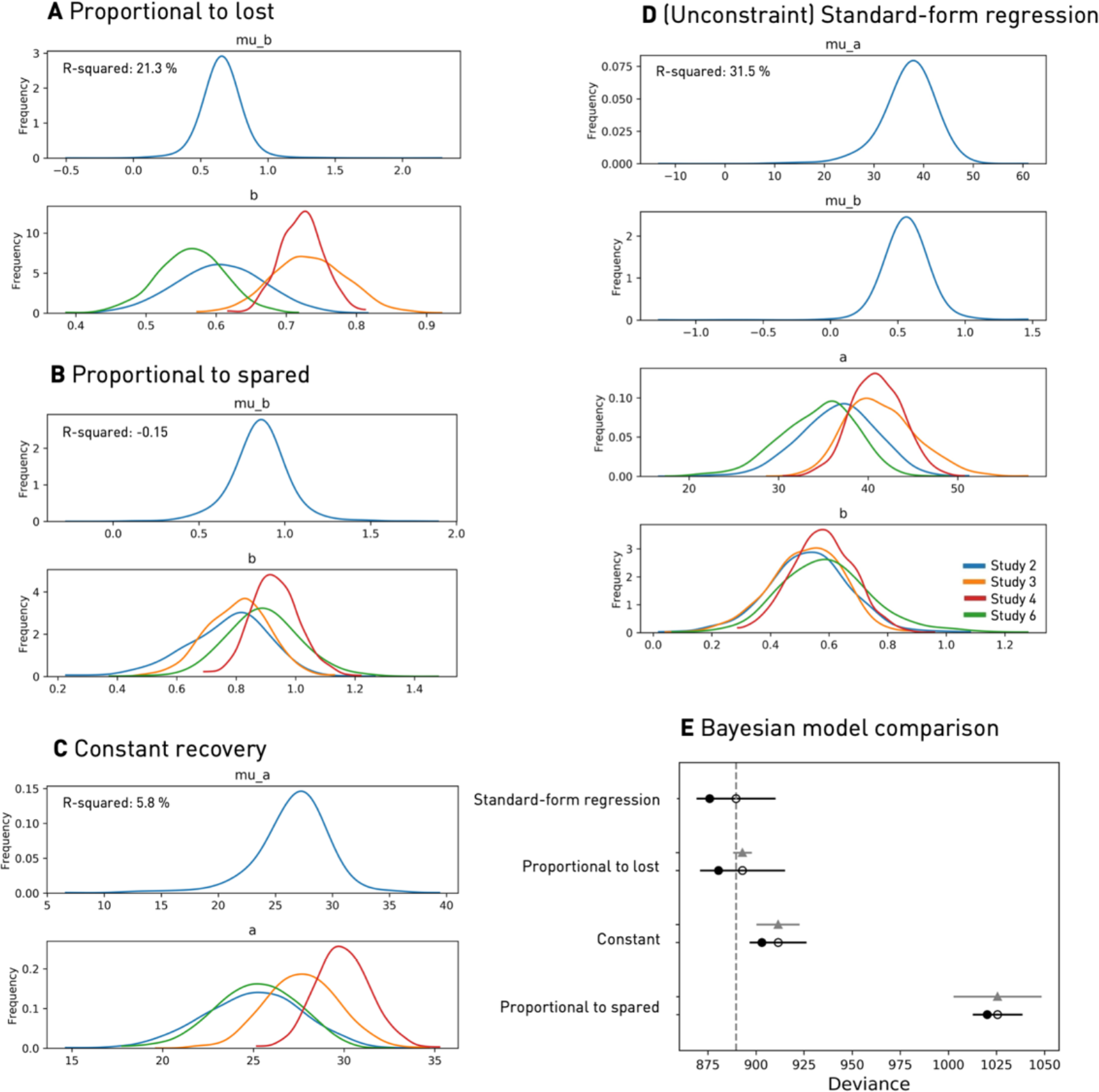
Alleviating confounds by ceiling effects and mathematical coupling: Bayesian hierarchical models in the subset of FM-initial 10-45. **Marginal posteriors for parameters of *proportional to lost function* recovery (A), *proportional to spared function* recovery (B), *constant* recovery (C), and *(Unconstrained) Standard-form* regression (D). E. Final Bayesian model comparison using leave-one-out-cross-validation (LOOCV)**. Empty circles represent the LOOCV-score, black error bars the corresponding standard error. The filled black circles mark the models’ in-sample deviances, grey triangles the difference to the top-ranked model as well as the associated standard error. Lastly, the lowest (i.e., best) LOOCV-value is indicated by the vertical dashed grey line. The standard-form regression model provides the best out-of-sample performance and is ranked first in the model comparison, closely followed by the *proportional to lost function* recovery model. Therefore, considering *fitters* only, recovery post-stroke seemed to follow a pattern resembling a mixture of *constant* and *proportional to lost function* recovery. However, the explained variance was low at 31.5% (standard-form regression) and 21.3% (*proportional to lost function* recovery).

The mean of the across-study slope-parameter in the *proportional to lost function* model equaled 0.65 (95% credibility interval 0.39–0.90), thus indicating an across-cohort recovery of a little less than 70%. In contrast to the model on the entire dataset, the explained variance came to just 21.3%. Notably, this value was lower than the explained variance based on the (unconstrained) standard-form regression model (PPC: R-squared: 31.5%). Across studies, subjects had a marginal posterior *constant* recovery of 26 points, ranging from 25 for Byblow *et al*. (2015) and Guggisberg *et al*. (2017) to 30 points for Winters *et al*. (2015). The explained variance amounted to only 5.8%. Explained variance dropped even further in case of *proportional to spared function* (PPC: R-squared: -0.153, slope=0.85), with the negative value signaling the unsuitability of this model. Since these fittings put us on the fringe of correct parameter retrieval for *constant* recovery and *proportional to spared*, we provide further justification for our conclusions in the supplementary materials section 9.

As the reported R-squared values are only comparable to a certain extent, since a model’s inherent degrees of freedom (i.e., flexibility) are not quantified in this measure, we performed a Bayesian model comparison based upon leave-one-out-cross-validated deviance values. The standard-form regression model, as well as the *proportional to lost* change model, had the lowest deviance and were thus top-ranked. The standard-form regression model gave a fit that can be seen as a combination of *constant* and *proportional to lost function* recovery and could thus be viewed as liberal as *proportional to lost*, c.f. **Supplementary Materials**. The *constant* recovery model followed these two models. *Proportional to spared function* performed the worst. Non-overlapping confidence intervals for the differences in deviance increased confidence in the two winning models (c.f. **Figure 7, E**; c.f. **Supplementary Figure 3** for WAIC-based results). Results for the additional subsets *FM-initial* 10–40 and 10–50 are broadly comparable to the subset *FM-initial* 10–45 (**Supplementary Figures 4 & 5)**.

When adjusting for missing values in our dataset in additional analysis, we determined the *upper bound* of the R-squared value for the winning, standard-form regression model to be 44.7%.

#### *Fitters* & *Non-Fitters* in the subset of FM-initial 0 – 45

Merging *non-fitters* and *fitters* increased the total sample size to 385 subjects, out of which 39 reached maximum values of 66 at follow-up (10.1%). Further characteristics, such as the ratio of standard deviations, are given in **Table 1**. Once again, we employed our *mitigating ceiling effect* approach and only considered subjects with *FM-initial* scores lower than 45, restricting our analysis to 270 subjects (out of 385, 70%; eight at absolute ceiling at follow-up, 3.0%). Posteriors of the various models’ parameters could be sampled reliably, as indicated by converging chains. Evaluating the standard-form regression model first: All of the individual slopes’ marginal posterior distributions had a mean in between 1.17 and 1.34, yet included 1 in their credibility intervals (across-cohort slope: 1.24; 95% credibility interval 0.99 – 1.52, c.f. **Figure 8, D**). Therefore, they indicated a mixture of *constant* and *proportional to spared function* recovery (PPC: R-squared: 52.8%), which could be related to the liberal proportional to spared discussed in the **Supplementary Material, section 1**. Two of the change models, i.e., *proportional to lost-function* and *proportional to spared function*, provided a very poor (negative) in-sample explained variance (PPC: R-squared: - 0.13 and – 0.51 for *proportional to lost* and *proportional to spared function* recovery, respectively, c.f. **Figure 8, A & B**). Only the *constant* recovery model could capture some positive variance (PPC: R-squared: 7.4%, c.f. **Figure 8, C**). The final model comparison revealed the (unconstrained) standard-form regression model, indicating a mixture of *constant* and *proportional-to-spared-function* recovery, and *constant* recovery as the winning models (c.f. **Figure 8, E**).

**Figure 8.**
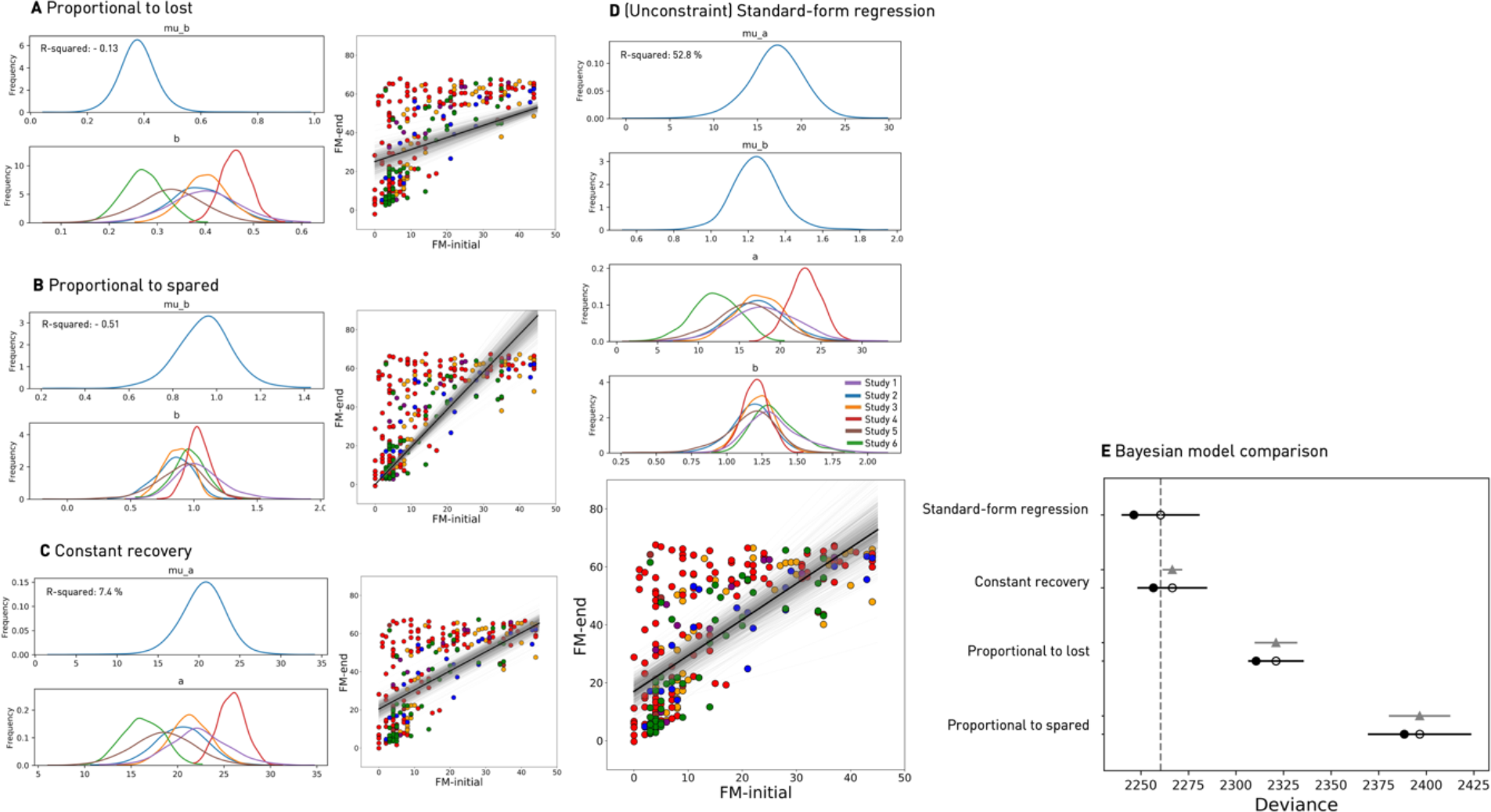
Joint hierarchical Bayesian analysis of *fitters* and *non-fitters* for FM-initial 0-45. A – D. Recovery models: p*roportional to lost function* recovery (A), *proportional to spared function* recovery (B), *constant* recovery (C), and *(Unconstrained) Standard-form* regression (D). Marginal posterior distributions are presented on the left-hand side for A – C and in the upper part of D. Distribution of initial against follow up scores in conjunction with an overlay of sampled fits are added on the right-hand sides for A – C and in the lower part of D (*thick black line:* mean, *grey lines:* 2000 sampled marginal posterior parameter constellations) **E. Final model comparison**. Based on leave-one-out-cross-validation, model comparison elected the standard-form regression model, indicating a mixture of *constant* and *proportional to spared function* recovery. Pure *constant* recovery was the best follow-up model.

## Discussion

Current analyses of proportional recovery after stroke are subject to various confounds. We here proposed a subset approach to minimize the key confound, *Compression enhanced Coupling*, which we validated in synthetic data experiments. We furthermore employed hierarchical Bayesian models to analyze the largest, compiled dataset of upper limb recovery post-stroke (n=385) and evaluate various conceivable patterns of stroke recovery in overall model comparisons.

In case of those patients considered *fitters* to the proportional recovery rule (*FM-initial*>10), model comparison pointed in the direction of either *proportional to lost function*, with a recovery proportion of 65% (i.e., slope of regression), or a combination of *constant* and *proportional to lost function* recovery as the underlying relationships. These findings were, therefore, generally in line with previous assumptions of *proportional to lost function* recovery post-stroke (Prabhakaran *et al*., 2008). However, and critically, the pure *proportional to lost function* recovery model could only explain 21% of the variance in recovery, a value drastically reduced in comparison to earlier studies, reporting up to 94% (Winters *et al*., 2015). Given the likely confounds by *Compression enhanced Coupling* in these earlier studies, the current estimate of explained variance may be considered more accurate, especially as it did not surpass the one for the classical standard-form regression – which we determined to be 32%. As the standard-form regression directly linked initial and follow-up FM scores, it is important to note that this model was not prone to the confounds due to coupling.

Of note, these conclusions substantially depended on the exclusion of *non-fitters*: a completely different picture arose when analyzing the entire spectrum of subjects, i.e., *fitters* (*FM-Initial*>10) and *non-fitters* (*FM-Initial*≤10) combined. The model comparison led to the election of a composite of *constant* and *proportional to spared function* recovery as the winning model^2^; the explained variance was 53%. In contrast to *proportional to lost function, proportional to spared function* recovery suggests that patients with greater preservation of function have a higher capacity to improve, which could have important implications for the conceptualization of recovery trajectories.

Eventually, the pressing questions are: Are we in need of more studies jointly analyzing *fitters* and *non-fitters*, particularly given the higher explained variance for the whole sample? Are 32% or even 53% explained variance sufficient to justify a recovery *rule* that may guide individual predictions in a future of precision neurology?

### Clinical importance and prediction of recovery post-stroke

Overestimation of the proportional recovery rule becomes particularly problematic when it impacts clinical practice, e.g., prompts the assumption of a spontaneous recovery process that exclusively depends on initial motor performance. In particular, such a conclusion may limit the allocation of valuable therapy sessions to some stroke subjects and generate a negative prior expectation towards tailored therapies (Byblow *et al*., 2015; Hawe *et al*., 2019*b*). Putting its suitability for single-participant prediction aside, Byblow and Stinear (2019) and Kundert *et al*. (2019) recently underscored the proportional recovery rule’s purpose for explaining the recovery process in stroke populations in general. From this perspective, it may be that *proportional to lost function* recovery explains *fitters’* trajectories better than other change models, but because of the level of explained variance, this does not make it, as such, a *good* explanation. Indeed, it is undoubtedly the case that a group-level effect size for a theory can be so small, that the theory has little explanatory value. Additionally, in either case, one should always strive to obtain estimates as un-confounded and accurate as possible.

Because of the increase in the explained variance of motor recovery when jointly considering *fitters* and *non-fitters* – from 32% to 53% – we may also need to rethink conventional analysis approaches and increasingly shift the focus on severely affected subjects by including *non-fitters* more often. This might be particularly important in view of substantial amounts of severely affected patients, e.g., 37% of *non-fitters* in our study and generally increasing numbers of patients with severe arm impairments (Hayward *et al*., 2017). Nevertheless, even 53% of explained variance leaves many unaccounted sources of variation in stroke recovery. This, for example, is evident in the visible heterogeneity of recoveries for patients with comparable initial scores; see the scatter plots in **Figure 3**. In this respect, our finding of higher explained variance when estimating parameters for *fitters* and *non-fitters* combined does not stand against the observation that there are grossly different recovery patterns, that may necessitate differing therapeutic (rehabilitative) approaches for variably impaired patient subgroups.

### Likert-like scales and ceiling effects

It is also essential to be aware of a particular score’s characteristics. The FM score is a Likert-like scale, thus a summary of multiple Likert-like items, comprising ordinal data (Likert, 1932). It is this combination of multiple items that renders parametric statistical approaches – that we applied here – feasible (Norman, 2010; Harpe, 2015). Positively, reported test-retest and inter-rater reliabilities for FM scores are *r>0*.*95* over repeated measurements (Gladstone *et al*., 2002). However, as previously emphasized, the FM assessment is highly susceptible to ceiling effects (Gladstone *et al*., 2002). We here further dissected these ceiling effects and highlighted the induction of *Compression enhanced Coupling* with change formulation models, that link initial scores and recovery (Lord, 1956; Hawe *et al*., 2019a; Hope *et al*., 2019). Amongst our total sample size of 385 subjects, 39 (10.1%) reached maximum scores at follow-up, and many more were likely compressed towards, but not at ceiling. Our synthetic data experiments showed that this degree of ceiling effect was sufficient to impede correct conclusions: Independent of the simulated ground truth mechanism, we would always discover *proportional to lost function* recovery.

We here relied upon the logic of subsets to decrease confounding effects by ceiling. That is, we focused on lower ranges of initial motor performance scores, which are less likely to lead to maximum follow-up motor performance scores. The relationships identified in the ceiling-reduced subsets could then be extended to the entire sample, as generalization was ensured by the assumed constant relationship between initial and follow-up scores. Such an assumption of constancy is inherent to linear regression models applied here. Importantly, our synthetic data experiments additionally demonstrated that we did not incur any new confounds, which would affect conclusions when defining subsets on initial scores. Altogether, we ensured more trustworthy model estimations by these means.

### Bayesian hierarchical models

Potentially insufficient numbers of subjects could endanger successful subset analyses relying on the data of just one study. This circumstance may be particularly so, as it may not be feasible to increase the size of individual studies, as high-quality data acquisition is time-consuming and costly. Here, the subset approach was only rendered possible due to our Bayesian hierarchical framework that facilitated the fusion of multiple datasets with individual-level data (Gelman and Hill, 2006). In this way, we could maximize the number of included subjects while retaining as much information on each study’s characteristics as possible and modeling uncertainty explicitly (McElreath, 2018). This combination of merging studies and preserving individual features was particularly appealing since it addressed both similarities and dissimilarities between individual studies, i.e., similar scores from similar patients at similar timepoints on the one hand and yet study sites and likely minor variations in therapies on the other hand. Also, our Bayesian hierarchical models were capable of effectively handling diverging sample sizes in the six studies considered (McElreath, 2018). Lastly, as anticipated in (Hope *et al*., 2019), they allowed for the evaluation of various generative models on the nature of recovery – *proportional to lost function, proportional to spared function*, and *constant recovery* – through model comparisons.

### Limitations and future directions

We decreased distorting ceiling effects by limiting analyses to subsets of initial scores. However, we acknowledge that there are drawbacks to this approach, such as the exclusion of substantial portions of the entire sample. Also, it does not represent definitive handling of the ceiling problem. The FM assessment is based on several single items (e.g. patient is able/partially able/unable to move his/her hand from the ipsilateral ear to the contralateral knee), which potentially results in multiple sub-ceilings. These may remain present even in case of discarding data at the scale’s maximum. Therefore, one viable strategy to circumvent these kinds of ceiling effects could be the increased use of continuous assessment scores, measuring, e.g., muscle forces, movement speeds or other kinematic parameters. Another strategy could be the construction of more elaborate scoring systems that allow for the detection of even very subtle variation, especially at the top of the scale. Nevertheless, motor performance may be maximally recovered and effectively indistinguishable from a healthy pre-stroke level in some cases. As a result, some natural ceiling would occur and require special attention, primarily concerning statistical proceedings. Future research may thus utilize our subset analysis or further approaches for censored data, such as (frequentist) Tobit models (Tobin, 1958) and Bayesian counterparts (Gelman and Hill, 2006).

Furthermore, we did not consider the possibility of any non-linearities of the FM scale, i.e., to differentiate between a ten-point gain from 0 to 10 and a ten point gain from 50 to 60. After all, the genuine underlying relationship of recovery may not be linear and might thus not be detectable by the linear regression models we, and the majority of the field, have so far focused on. Therefore, our results encourage research into other model types, for example, non-linear models, such as decision tree-like algorithms (Stinear *et al*., 2012). Additionally, we may need to refocus on a variety and multivariate combination of indicators of stroke recovery, such as behavioral, physiological, and imaging biomarkers, which have already shown promise (Stinear, 2017; Ward, 2017; Findlater *et al*., 2019).

Our study highlights the opportunity for novel insights to be gleaned by Bayesian hierarchical modeling, as it facilitates model comparisons and the creation of large datasets, thereby increasing the generalizability of attained inferences. Therefore, they are likely to become common in stroke research, as well as in other clinical fields. Indeed, the strategies outlined here may inspire and guide future studies, raise awareness of the better handling of ceiling and change models, as well as the pernicious nature of *compression enhanced coupling*; especially, as these effects may frequently occur in biomedical data.

In this present study, we relied on an already large number of 385 subjects. Nonetheless, we are still in need of larger stroke recovery datasets that additionally capture a multitude of stroke symptoms, such as motor symptoms of upper and lower limbs, aphasia, neglect and hemianopia. In particular, the field needs to take advantage of collaborative data collection and move beyond behavioral scores for both predictor and outcome variables. In this way, a range of potential explanatory and predictive variables could be incorporated to reliably increase explained variance and accuracy of out-of-sample prediction of stroke recovery - in case of *fitters* and also *non-fitters (*Grefkes and Fink, 2016; Boyd *et al*., 2017; Bernhardt *et al*., 2017).

### Conclusion

Our probabilistic generative analyses of post-stroke motor performance in a sample of 385 stroke participants revealed only weak signs of *proportional to lost function* recovery for those defined to be *fitters* to the proportional recovery rule. Variance in recovery could be explained by up to 32% only, more than 50% less than previously reported. Additionally, a combination of *constant* and *proportional to spared function* emerged as a likely relationship for the recovery of the entirety of stroke subjects – at a higher explained variance of 53%. Importantly, these estimates were obtained after de-confounding effects of mathematical coupling and ceiling (Hawe *et al*., 2019a; Hope *et al*., 2019). In summary, these lower levels of explained variance may motivate research moving beyond behavioral measures and the consideration of combinations of various biomarkers, such as demographical, clinical, imaging, and physiological ones. Ultimately, our findings may also pave the way for more frequent use of Bayesian hierarchical analyses to distill conclusions resting upon merged clinical datasets, efficiently ensuring reliable generalization performance and modelling of uncertainty.

## Data Availability

The recovery data as well as jupyter notebooks (python 3.7, primarily software package pymc3) employed in this study are available from the authors on reasonable request.

## Acknowledgements

We thank Michael Moutoussis for valuable observations and discussions.

## Abbreviations

CeC: Compression enhanced coupling
FM: Fugl Meyer
PPC: Posterior predictive check

## Supplemental materials

### 1 Transformation of *change* equations (c.f. Introduction Section of Main Body)

We show that all the *change* equations can be re-expressed as simple direct linear regression, what we call standard-form regression, bringing them all into the same model framework.

This mathematical formulation is inspired by the Fugl-Meyer (FM) scale for assessing upper limb deficits post-stroke (Prabhakaran et al., 2008). Specifically, *X*∈ ℝ^0,*Max*^ denotes performance scores at time point 1 (i.e., *FM-initial* in the main body of the paper), *Y*∈ ℝ^0,*Max*^ denotes performance at time point 2 (i.e., *FM-end*), and *Max*∈ ℝ^7^is the top of the scale, which corresponds to “normal” performance on the scale (66 in the FM scale).

We begin by defining the basic change equations, so-called because the dependent variable is the change from time-point 1 to time-point 2 (i.e., recovery). For simplicity of presentation, we do not include noise terms, but these could be added in the usual way.

**Definitions**

1. Classic *proportional to lost* function is defined as,

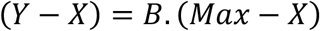

where *B*∈ ℝ ∧ 0 ≤ *B*<1.
2. *Proportional to spared* function is defined as,

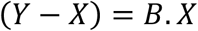

where *B*∈ ℝ ∧ 0 ≤ *B*<1.
3. *Constant* is defined as,

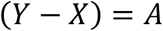

where *A* ∈ ℝ ∧ 0 ≤*A*< *Max*. ∎

These three change formulae generate different patterns in all but one case, which is when *B*= *A*=0, the boundary condition in which *Y*= *X*and there is no recovery.

The standard-form linear regression equation, which directly relates *Y*and *X*, rather than through change, is defined as,

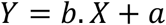

where *a, b* ∈ ℝ. We use lower case to distinguish coefficient and intercept when associated with standard-form regression and upper case when associated with the change formulations.

We now show that each change formulation can be rearranged into a standard-form regression.

**Proposition 1 (Proportional to Lost)**

Given that 0 ≤ *B*<1,

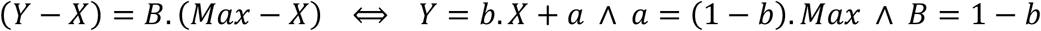

**Proof**

Assume 0 ≤ *B*<1, then we can reason as follows,

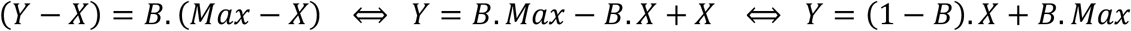

but now, if we let *b* =1 − *B*and *a*=*B*.*Max*, we obtain,

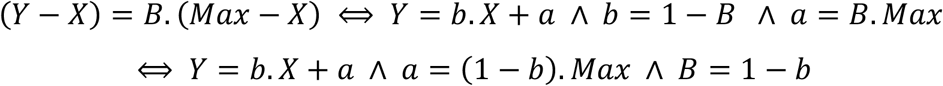

which is as required. ∎

Proposition 1 shows that once the proportional to lost equation is turned into the standard-form regression equation, as *a* goes up (in the range zero to *Max*), *b* comes down, according to the formula *b* =1 − (*a*/*Max*). Thus, stronger proportional recovery patterns, i.e., with bigger *B*’s, lead to increased intercepts and reduced slopes in the standard-form regression.

The set of possible linear relationships that proportional to lost encompasses are shown in **figure 2** (panel A change form and D standard-form) in the main body. This makes it clear that proportional to lost reflects a restricted range of the standard-form regression. Indeed, we can highlight two varieties of proportional to lost, both of which are sub-models of standard-form regression:

a. *Liberal proportional to lost*: standard-form regression in which 0 < *b* ≤1 and 0 ≤*a*< *Max*. This generalisation enables, for example, constant recovery to be mixed with strong proportional to lost.
b. *Strong proportional to lost*: the classic change formulation of proportional to lost, which (putting mathematical coupling aside) is equivalent to standard-form regression with 0 < *b* ≤1 and *a*=(1− *b*).*Max*.

Both these forms require the slope to be less than or equal to the identity function (i.e., *b* ≤1). *Liberal* proportional to lost, though, does not constrain the relationship between slope and intercept, while *strong* proportional to lost does, making *strong* a sub-model of *liberal*.

Notably, the liberal and the strong proportional to lost formulations behave differently with regard to ceiling. That is, it is easy to show that, in the absence of noise, strong proportional to lost cannot generate a ceiling effect, i.e., *Y*values above *Max*. Specifically, strong proportional to lost can be expressed as,

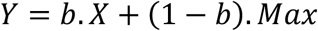

where, for a given *b*, the second summand on the right is constant and the first summand is maximal when *X*= *Max*. Thus, the maximum *Y*value that can be obtained is *b*.*Max*+ (1− *b*).*Max*, which equals *Max*. In contrast, since *b* and *a* are independent in liberal proportional to lost, both can be set high in their given range, generating *Y*values above *Max*. Now, we move to consider proportional to spared function.

**Proposition 2 (Proportional to Spared)**

Given that 0 ≤ *B*<1,

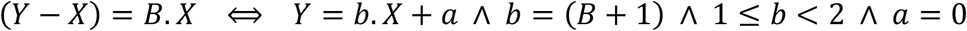

**Proof**

Straightforward. ∎

So, proposition 2 implies that proportional to spared can be expressed in standard-form regression, with an intercept of zero and a slope that is steeper than or equal to the identity, but less than 2. This range of possible linear relationships is depicted in **Figure 2** (panel B change formulation and E standard-form).

As was the case for proportional to lost, we can also identify a liberal form of proportional to spared.

a. *Liberal proportional to spared*: standard-form regression in which 1≤ *b* < 2and 0 ≤*a*< *Max*.
b. *Strong proportional to spared*: the classic change formulation of proportional to spared, which (apart from mathematical coupling) is equivalent to standard-form regression with 1≤ *b* < 2and *a*=0.

Both these forms require the slope to be greater than or equal to the identity function, but less than 2. *Strong* proportional to spared, though, does not allow the intercept to increase above zero, while liberal proportional to spared does, making *strong* a sub-model of *liberal*.

As shown in **Figure 2** (panel E), strong proportional to spared generates ceiling effects, even without adding noise once *b*>1 (i.e., once *B*>0). These ceiling effects increase in severity as *b* (or, indeed, *B*) gets bigger. Ceiling effects become even more severe if the intercept is allowed to be greater than zero, as supported by liberal proportional to spared.

Finally, we consider constant recovery (which is, of course, different to constant for standard-form regression, which would be *Y*=*a*).

**Proposition 3 (Constant Recovery)**

Given that 0 ≤*A*< *Max*,

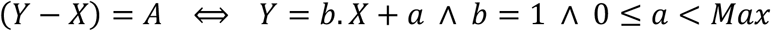

**Proof**

Straightforward. ∎

This reformulation makes clear that constant recovery generates the range of possible linear relationships shown in **Figure 2** (panels C and F). For *a* bigger than zero, it also generates increasingly severe ceiling effects as *a* increases, even without a noise term.

### 2 Summary of Model Formulations (c.f. Introduction Section of Main Body)

We employ several different model formulations in this paper. We summarise these formulations and where they are used here.

**Ground Truth Simulations (c**.**f. subsection “Synthetic data simulation experiments” in “Results” section)**

We generate fake, i.e., synthetic, data according to (constrained) standard-form regression models, of the form,

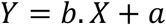

where the constraints are exactly as in **figure 2**, i.e., proportional to lost (panel D), proportional to spared (panel E), and constant recovery (panel F).

**Fitted to Ground Truth Simulation Data (c**.**f. subsection “Synthetic data simulation experiments” in “Results” section)**

In all cases, we fit the model

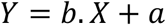

to the ground-truth simulation data; so we obtain estimates of slope, *b*, and intercept, *a*, (along with their confidence intervals). Then, we assess the accuracy of fitting by comparing to the true slope and intercept; c.f. previous section.

**Fitted to Human Data, before Discarding Ceiling (c**.**f. subsection “Bayesian hierarchical model” in “Results” section and Figure 4)**

This is the first fitting to human data. We fit two models:

1. (unconstrained) standard-form regression, i.e., *Y*= *b*.*X*+*a*.
2. Typical change-form regression, i.e., *Y*− *X*= *b*.(*Max*−*X*) +*a*.

To fully illustrate how extreme the effect of Compression Enhanced Coupling can be on the fitting of the change form model, we allowed corresponding flexibility to the change- and standard-form equations and incorporated an intercept (*a*) into the change-form model.

**Fitted to Human Data, with Ceiling Discarded (c**.**f. subsection “Final model comparison (on human data) in the subset of FM-initial 10-45” of “Results” section)**

We fitted the following models to the human data.

1. (unconstrained) standard-form regression, i.e., *Y*= *B*.*X*+ *A*.
2. Change-form proportional to lost, i.e., *Y*− *X*= *B*.(*Max*−*X*).
3. Change-form proportional to spared, i.e., *Y*− *X*= *B*.*X*.
4. Change-form constant recovery, i.e., *Y*− *X*= *A*.

2, 3, and 4, here, are as per panels A, B, and C of **Figure 2**.

## 3 Data acquisition and details on the Fugl Meyer Scale (c.f. section Methods of Main Body)

We contacted nine corresponding authors of eleven studies comprising longitudinal data on motor function measured on the Fugl Meyer scale post-stroke (screening of titles & abstracts, keywords “poststroke”, “recovery”, “motor function”, “longitudinal”, “Fugl-Meyer” on PubMed as well as following referenced literature, criteria: > 20 stroke patients, initial and follow-up Fugl Meyer Score of the upper limb; initial: acute phase post-stroke, follow-up: 3-6 months post-stroke). The response rate was 56 %. Only two authors ((Hawe, Scott, & Dukelow, 2019), (Guggisberg, Nicolo, Cohen, Schnider, & Buch, 2017)) were able to share their data (reasons for the decline: limited by ethics, in-house study). As individual-level data from (Zarahn et al., 2011) was openly available, we were, therefore, able to merge it with data from (Hawe et al., 2019) and (Guggisberg et al., 2017). Please see (Hawe et al., 2019) for a more detailed description of the data extraction. In brief, published figures of initial and end FM scores were digitized and positions of points extracted in Matlab, missing data was 17.7 %, primarily due to overlapping data points.

The FM assessment is a quantitative instrument to determine sensorimotor impairment post-stroke, divided into five domains (motor function, sensory function, balance, joint range of motion, and joint pain) (Fugl-Meyer *et al*., 1975). Please note that we considered information on the motor performance of the affected upper limb only; a minimum of 0 implied no preserved and a maximum of 66 indicated full motor function of the upper limb (multiple items on movement, coordination, reflex action on the shoulder, elbow, forearm, wrist, hand, each evaluated on a 3-point ordinal scale (0 = no performance, 1 = partial performance, 2 = full performance).

## 4 Defining simulated ceiling: Details on measurement noise (c.f. section Methods of Main Body)

When simulating the outcome, *FM-end*, we first added “State noise” (Gaussian noise, mean = 0, Standard deviation (SD) = 5) to the output of the linear ground truth model. “State noise” can be understood as unexplained randomness associated with the internal state of the system, e.g., variation in the alertness of subjects. This form of noise reflects the actual capacity of the brain, ignoring the measurement method, and thus, naturally, is not bounded by the Fugl-Meyer scale. Next, we enforced ceiling by setting any value greater than 66 to 66, i.e., the maximum value of the Fugl Meyer assessment. This imposition of ceiling reflects the application of the Fugl-Meyer scale and the parameters associated with it. This testing process will naturally involve a second source of random variability – so-called measurement noise. The latter reflects variability in the administration of the scale, which can be as subtle as the motivation a tester engenders in the patient and him- or herself. We justify our setting of measurement noise (Gaussian noise, mean = 0, SD = 2) in what follows. Since this further application of Gaussian noise will place some data points above ceiling, we enforced a final ceiling by once again setting any value greater than 66 to 66.

We relied on information about the test-retest reliability of the FM-scale, when estimating an appropriate level of measurement noise. Reported test-retest and inter-rater reliabilities for the Fugl Meyer are *r>=0*.*96* over repeated tests (Gladstone, Danells, & Black, 2002). Accordingly, we assessed the ratio of the data SD that the noise SD has to be to obtain such an *r*-value. This correlation is between a first and second test, where the only difference between the two is the addition of the noise.

The results of this simulation are shown in **Supplementary Figure 1**, which demonstrates that noise with 10-20% (or even up to 30%) of the standard deviation of the original scores, gives strong enough correlations to match empirically reported *r-values*. Given our “true” models, standard deviations of the outcome had a range of 5.3 – 33.7. For reasons of simplicity, we calculated a probable level of noise based on the smallest occurring standard deviation, i.e., SD = 5.3, and an assumption of 30 % noise for all models; this gives us a rounded measurement noise standard deviation value of 2.

**Supplementary Figure 1.**
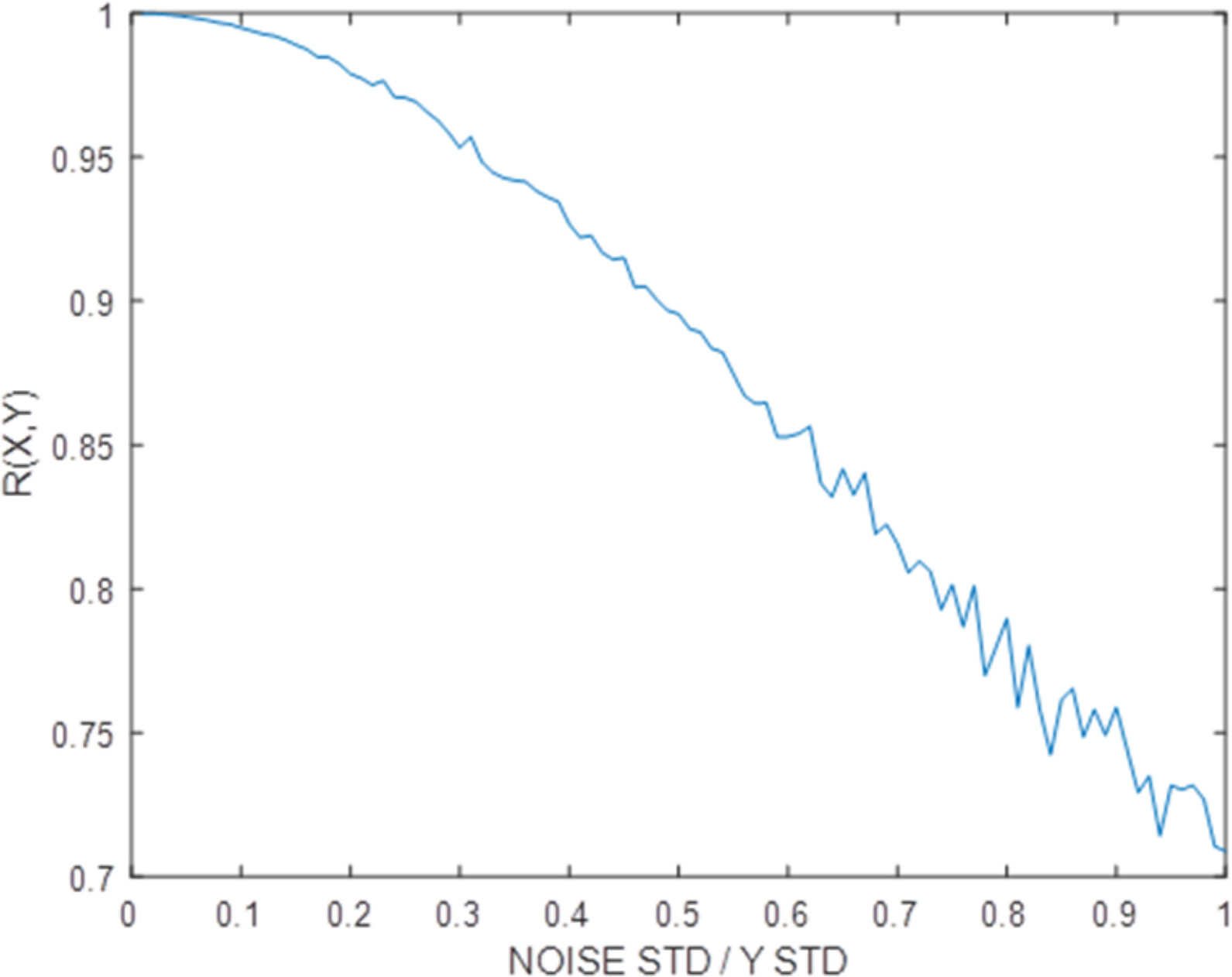
Correlation of variables X and Y, representing two measurements, where X is Y plus zero mean Gaussian noise. The standard deviation of the Gaussian noise is varied relative to a fixed standard deviation for X.

## 5 Additional comments on simulations (c.f. section Methods of Main Body)

Importantly, the model comparison problem we are undertaking is simplified because the linear fits provided by each of the three recovery theories – proportional to lost, to spared, and constant – are, in all but one case, non-overlapping. This overlap can be seen in panels [A. B & C], as well as [D, E & F] of **Figure 2**. Specifically, whether in change or standard-form, apart from the identity mapping, i.e., Y = X (which all the models allow, but will not arise in our fitting), the model spaces are disjoint. This effectively ensures *model* retrievability, i.e., if we can show that each model can retrieve the parameters of synthetic data it generated (the issue considered in section 8 of the supplementary material), no non-generating model will fit better than a generating model.

Of course, this only applies to the three specific recovery models – proportional to lost, proportional to spared, and constant recovery. We will also fit (unconstrained) standard-form regression, which can generate all the linear fits of the three specific models, as well as fits that they cannot generate.

## 6 Bayesian hierarchical models for the standard-form regression and change model of *fitters* (*FM-initial*: > 10): Posterior predictive checks (c.f. Results section of Main Body)

**Supplementary Figure 2.**
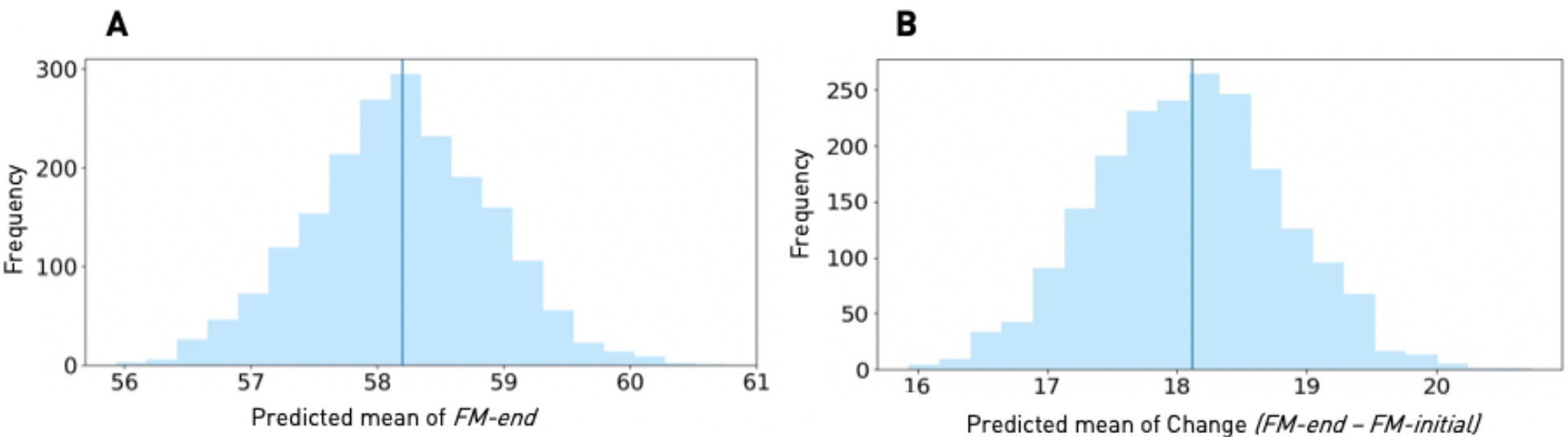
Bayesian hierarchical models: Posterior predictive checks. **A** (Unconstrained) Standard-form regression. **B** Change model. Histograms of the predicted means of *FM-end* in case of the standard-form regression or *Change* in case of the change model based on the sampled posteriors in comparison to the true mean of *FM-end* or *Change* (darker blue vertical line). Agreement of means indicates reliable predictions of patterns that are present in the real data.

## 7 Detailed evaluation of simulations (c.f. Results section of Main Body)

We report here the results of our ground truth simulations, which are performed for the three key models: *proportional to lost function recovery, proportional to spared function recovery*, and *constant recovery*. In each of these cases, data is generated according to the standard-form regression version of each model, i.e., the equations presented in panels [D, E, and F] of **Figure 2**. These re-expressions are justified in supplementary materials 1. Thus, the generating models can be expressed as *Y=b*.*X+a*, with *b* the true slope and *a* the true intercept in the tables. Also, for proportional to lost function recovery *b=1-B* and *a=B*.*Max*, with the “amount of recovery” (1^st^ column of tables) equal to *B*.*100*; for proportional to spared function recovery *b=(1+B)* and *a=0*, with “amount of recovery” equal to *B*.*100*; and for constant recovery *b=1* and *a=A*, with “amount of recovery” equal to *A*.

Each row in each table below corresponds to simulations in which we generate 1000 data sets according to given ground truth (i.e., setting of *b* and *a*) and then fit the same model (*Y=b*.*X+a*) to each data set. The critical criterion for judging the effectiveness of model retrieval is the number of times the true parameter settings (i.e., for *a* and *b*) were in the confidence intervals resulting from model fitting; we report the proportion that met the 68% and 95% Confidence Intervals (CIs). Model retrieval is maximally accurate if 68% (respectively 95%) of fits are in the 68% (respectively 95%) CIs. We also report the variability ratio, i.e., the ratio of standard deviations of *Y* and *X* (log of this number is shown in **Figure 1**) and then the two relevant correlations: *r(X,Y)* and *r(X,Y-X*), i.e., *FM-initial – FM-end and FM-initial – Change*. Finally, we report the average number of subjects at or above absolute ceiling, i.e., the *Y≥Max*=*66*.

We first state a summary of simulation results and then present the corresponding tables:

### Proportional to lost function recovery

#### Baseline models (state noise only)

Proportional to lost function recovery in the range of 10 % to 90 % was characterized by positive intercepts and slopes in-between values of 0 and 1. While intercepts increased with the amount of proportional to lost function recovery (range: 6.6 – 59.4), the opposite behavior was found for slopes (range: 0.9 – 0.2), Supplementary Table 1a illustrates these trajectories. This is consistent with proportional to lost function recovery when formulated as a standard-form regression; see **proposition 1** in supplementary materials (part 1) and **figure 2** panel [D]. Additionally, as expected and highlighted in (Hope *et al*., 2018) and (Hawe *et al*., 2019*a*), Pearson correlation values steadily decreased for *FM-initial – Change* until an almost perfect anti-correlation of -1, which was contrasted with decreasing positive correlation values for *FM-end – FM-initial*, when increasing absolute percentages of proportional to lost function recovery. Further, higher percentages of proportional to lost function were associated with a decrease in standard deviation ratio, reaching a minimum value of 0.3 for 90% recovery (c.f. **Figure 6, B** and C, left plots). In case of 70 % recovery, the absolute value of the correlation of *FM-initial – Change* surpassed *FM-end – FM-initial* by 0.24, therefore, perfectly demonstrating the inflation of correlation due to mathematical coupling and its enhancement as the variability ratio reduces.

#### Ceiling enforced

Enforcing ceiling aggravated the dissociation of correlations of *FM-initial – Change* and *FM-end – FM-initial*, in line with predictions by (Hope *et al*., 2018). The percentage of subjects at absolute ceiling (i.e. FM-end = 66) reached a maximum of 21.4 % for 90 % recovery (range: 4.1 % - 21.4 %, 70 % proportional recovery: 14.8 %).

#### Subset approaches FM-initial 10 to 60, 50 and 40

Reducing the number of subjects according to their *FM-initial* scores re-established accurate inferences of intercepts and slopes, as was indicated by met confidence intervals. The most substantial subset *FM-initial* 10 to 60 enabled veridical model estimation up to 30 % of proportional to lost function recovery, *FM-initial* 10 to 50 up to 70 % and *FM-initial* 10 to 40 for the entirety of models. This increase in accurate model estimation was paralleled by a decrease in subjects at ceiling.

**Supplementary table 1a:** Proportional to lost function recovery: No ceiling enforced.

| Amount of recovery | True Intercept | Estimated intercept | Met 68%-CI | Met 95%-CI | True Slope | Estimated slope | Met 68%-CI | Met 95%-CI |
| --- | --- | --- | --- | --- | --- | --- | --- | --- |
| 10% | 6.60 | 6.61 | 0.69 | 0.95 | 0.90 | 0.90 | 0.70 | 0.94 |
| 20% | 13.20 | 13.21 | 0.68 | 0.95 | 0.80 | 0.80 | 0.71 | 0.96 |
| 30% | 19.80 | 19.81 | 0.69 | 0.95 | 0.70 | 0.70 | 0.69 | 0.96 |
| 40% | 26.40 | 26.38 | 0.68 | 0.96 | 0.60 | 0.60 | 0.68 | 0.94 |
| 50% | 33.00 | 32.94 | 0.70 | 0.96 | 0.50 | 0.50 | 0.68 | 0.96 |
| 60% | 39.60 | 39.57 | 0.68 | 0.95 | 0.40 | 0.40 | 0.70 | 0.94 |
| 70% | 46.20 | 46.20 | 0.67 | 0.94 | 0.30 | 0.30 | 0.69 | 0.92 |
| 80% | 52.80 | 52.83 | 0.69 | 0.95 | 0.20 | 0.20 | 0.69 | 0.95 |
| 90% | 59.40 | 59.37 | 0.71 | 0.94 | 0.10 | 0.10 | 0.68 | 0.95 |

| **Ratio STD**<br>**Y/X** | **Correlation**<br>**X - Y** | **Correlation**<br>**X - Change** |
| --- | --- | --- |
| 0.94 | 0.95 | -0.33 |
| 0.85 | 0.94 | -0.57 |
| 0.76 | 0.93 | -0.72 |
| 0.66 | 0.90 | -0.81 |
| 0.58 | 0.87 | -0.87 |
| 0.49 | 0.82 | -0.90 |
| 0.41 | 0.72 | -0.93 |
| 0.35 | 0.57 | -0.94 |
| 0.30 | 0.33 | -0.95 |

**Supplementary table 1b:** Proportional to lost function recovery: Ceiling enforced.

| **Amount of**<br>**recovery** | **True**<br>**Intercept** | **Estimated**<br>**intercept** | **Met**<br>**68%-CI** | **Met**<br>**95%-CI** | **True**<br>**Slope** | **Estimated**<br>**slope** | **Met**<br>**68%-**<br>**CI** | **Met**<br>**95%-**<br>**CI** |
| --- | --- | --- | --- | --- | --- | --- | --- | --- |
| 10% | 6.60 | 7.09 | 0.61 | 0.90 | 0.90 | 0.88 | 0.49 | 0.85 |
| 20% | 13.20 | 13.72 | 0.59 | 0.90 | 0.80 | 0.78 | 0.46 | 0.83 |
| 30% | 19.80 | 20.38 | 0.56 | 0.87 | 0.70 | 0.68 | 0.43 | 0.80 |
| 40% | 26.40 | 27.09 | 0.51 | 0.83 | 0.60 | 0.57 | 0.35 | 0.70 |
| 50% | 33.00 | 33.71 | 0.51 | 0.85 | 0.50 | 0.47 | 0.27 | 0.68 |
| 60% | 39.60 | 40.37 | 0.47 | 0.82 | 0.40 | 0.37 | 0.22 | 0.55 |
| 70% | 46.20 | 47.01 | 0.42 | 0.79 | 0.30 | 0.26 | 0.14 | 0.44 |
| 80% | 52.80 | 53.56 | 0.42 | 0.81 | 0.20 | 0.16 | 0.06 | 0.28 |
| 90% | 59.40 | 59.39 | 0.61 | 0.92 | 0.10 | 0.07 | 0.08 | 0.33 |

| **Ratio STD**<br>**Y/X** | **Correlation**<br>**X - Y** | **Correlation**<br>**X - Change** | **Numbers of subjects at or**<br>**above absolute ceiling** |
| --- | --- | --- | --- |
| 0.93 | 0.95 | -0.37 | 10 |
| 0.83 | 0.94 | -0.60 | 11 |
| 0.74 | 0.92 | -0.74 | 13 |
| 0.64 | 0.90 | -0.83 | 14 |
| 0.55 | 0.86 | -0.88 | 17 |
| 0.46 | 0.80 | -0.92 | 21 |
| 0.38 | 0.70 | -0.94 | 26 |
| 0.30 | 0.53 | -0.96 | 35 |
| 0.23 | 0.29 | -0.97 | 52 |

**Supplementary table 1c:**
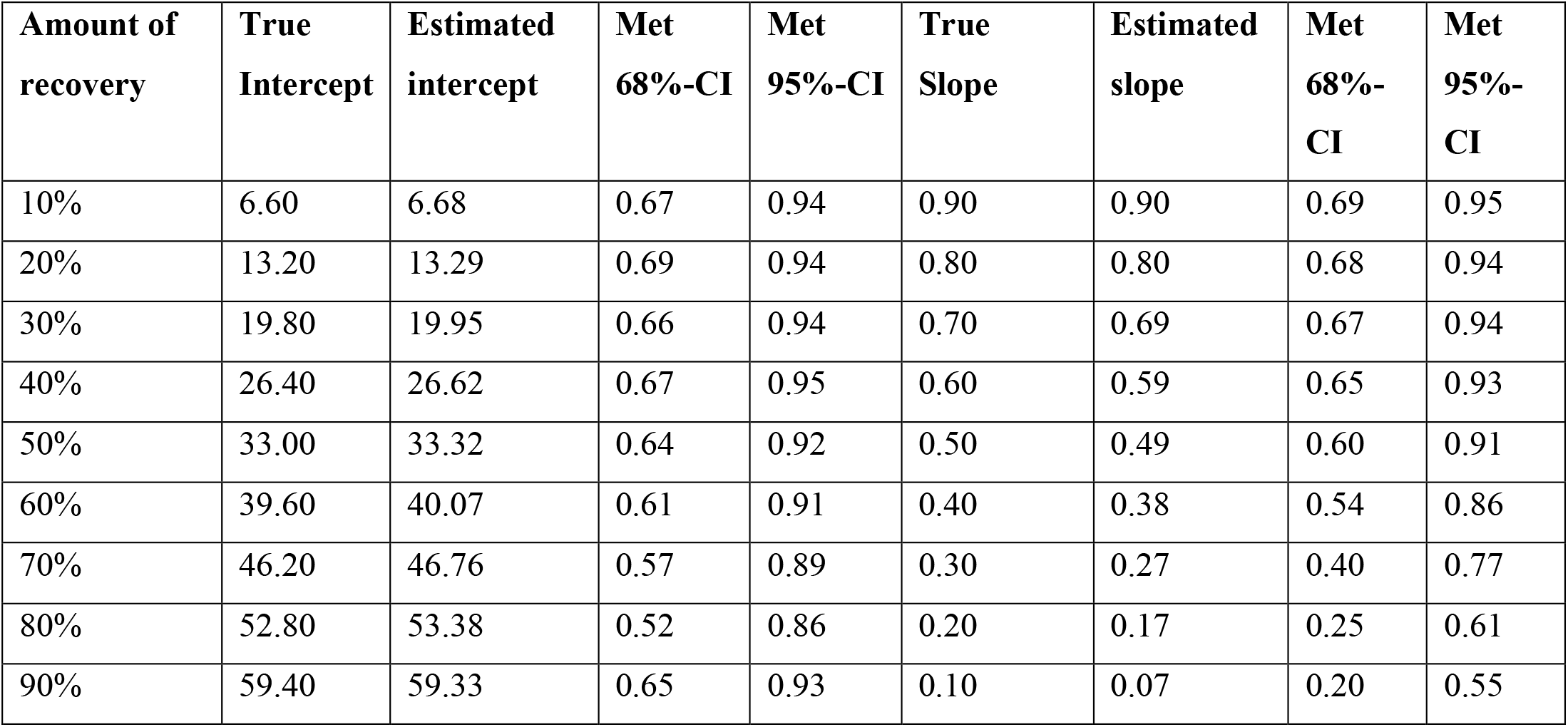
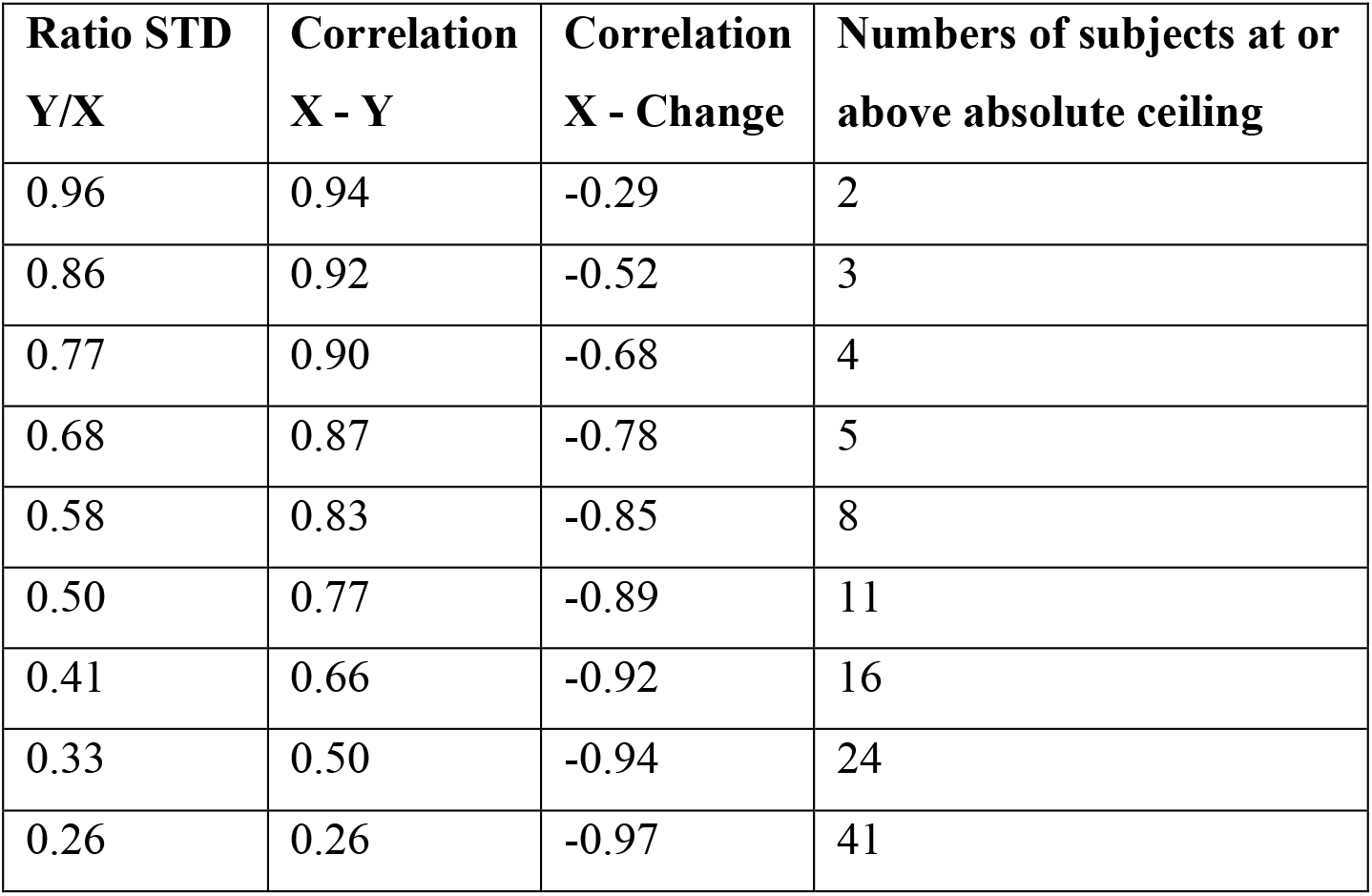
Proportional to lost function recovery: Ceiling enforced, Subset 10 - 60.

| **Amount of recovery** | **True Intercept** | **Estimated intercept** | **Met 68%-CI** | **Met 95%-CI** | **True Slope** | **Estimated slope** | **Met 68%-CI** | **Met 95%-CI** |
| --- | --- | --- | --- | --- | --- | --- | --- | --- |
| 10% | 6.60 | 6.68 | 0.67 | 0.94 | 0.90 | 0.90 | 0.69 | 0.95 |
| 20% | 13.20 | 13.29 | 0.69 | 0.94 | 0.80 | 0.80 | 0.68 | 0.94 |
| 30% | 19.80 | 19.95 | 0.66 | 0.94 | 0.70 | 0.69 | 0.67 | 0.94 |
| 40% | 26.40 | 26.62 | 0.67 | 0.95 | 0.60 | 0.59 | 0.65 | 0.93 |
| 50% | 33.00 | 33.32 | 0.64 | 0.92 | 0.50 | 0.49 | 0.60 | 0.91 |
| 60% | 39.60 | 40.07 | 0.61 | 0.91 | 0.40 | 0.38 | 0.54 | 0.86 |
| 70% | 46.20 | 46.76 | 0.57 | 0.89 | 0.30 | 0.27 | 0.40 | 0.77 |
| 80% | 52.80 | 53.38 | 0.52 | 0.86 | 0.20 | 0.17 | 0.25 | 0.61 |
| 90% | 59.40 | 59.33 | 0.65 | 0.93 | 0.10 | 0.07 | 0.20 | 0.55 |

| **Ratio STD Y/X** | **Correlation X - Y** | **Correlation X - Change** | **Numbers of subjects at or above absolute ceiling** |
| --- | --- | --- | --- |
| 0.96 | 0.94 | -0.29 | 2 |
| 0.86 | 0.92 | -0.52 | 3 |
| 0.77 | 0.90 | -0.68 | 4 |
| 0.68 | 0.87 | -0.78 | 5 |
| 0.58 | 0.83 | -0.85 | 8 |
| 0.50 | 0.77 | -0.89 | 11 |
| 0.41 | 0.66 | -0.92 | 16 |
| 0.33 | 0.50 | -0.94 | 24 |
| 0.26 | 0.26 | -0.97 | 41 |

**Supplementary table 1d:** Proportional to lost function recovery: Ceiling enforced, Subset 10 - 50.

| Amount of recovery | True Intercept | Estimated intercept | Met 68%-CI | Met 95%-CI | True Slope | Estimated slope | Met 68%-CI | Met 95%-CI |
| --- | --- | --- | --- | --- | --- | --- | --- | --- |
| 10% | 6.60 | 6.64 | 0.66 | 0.95 | 0.90 | 0.90 | 0.67 | 0.95 |
| 20% | 13.20 | 13.14 | 0.69 | 0.95 | 0.80 | 0.80 | 0.69 | 0.94 |
| 30% | 19.80 | 19.86 | 0.69 | 0.96 | 0.70 | 0.70 | 0.70 | 0.96 |
| 40% | 26.40 | 26.42 | 0.65 | 0.94 | 0.60 | 0.60 | 0.67 | 0.95 |
| 50% | 33.00 | 33.02 | 0.67 | 0.94 | 0.50 | 0.50 | 0.67 | 0.95 |
| 60% | 39.60 | 39.78 | 0.69 | 0.95 | 0.40 | 0.39 | 0.68 | 0.95 |
| 70% | 46.20 | 46.43 | 0.65 | 0.95 | 0.30 | 0.29 | 0.69 | 0.95 |
| 80% | 52.80 | 53.13 | 0.62 | 0.92 | 0.20 | 0.18 | 0.55 | 0.90 |
| 90% | 59.40 | 59.27 | 0.67 | 0.94 | 0.10 | 0.07 | 0.46 | 0.81 |

| Ratio STD Y/X | Correlation X - Y | Correlation X - Change | Numbers of subjects at or above absolute ceiling |
| --- | --- | --- | --- |
| 0.99 | 0.90 | -0.23 | 0 |
| 0.91 | 0.88 | -0.42 | 0 |
| 0.82 | 0.85 | -0.58 | 0 |
| 0.73 | 0.82 | -0.69 | 1 |
| 0.65 | 0.76 | -0.76 | 1 |
| 0.57 | 0.68 | -0.82 | 3 |
| 0.50 | 0.58 | -0.87 | 6 |
| 0.42 | 0.42 | -0.91 | 12 |
| 0.34 | 0.21 | -0.94 | 27 |

**Supplementary table 1e:**
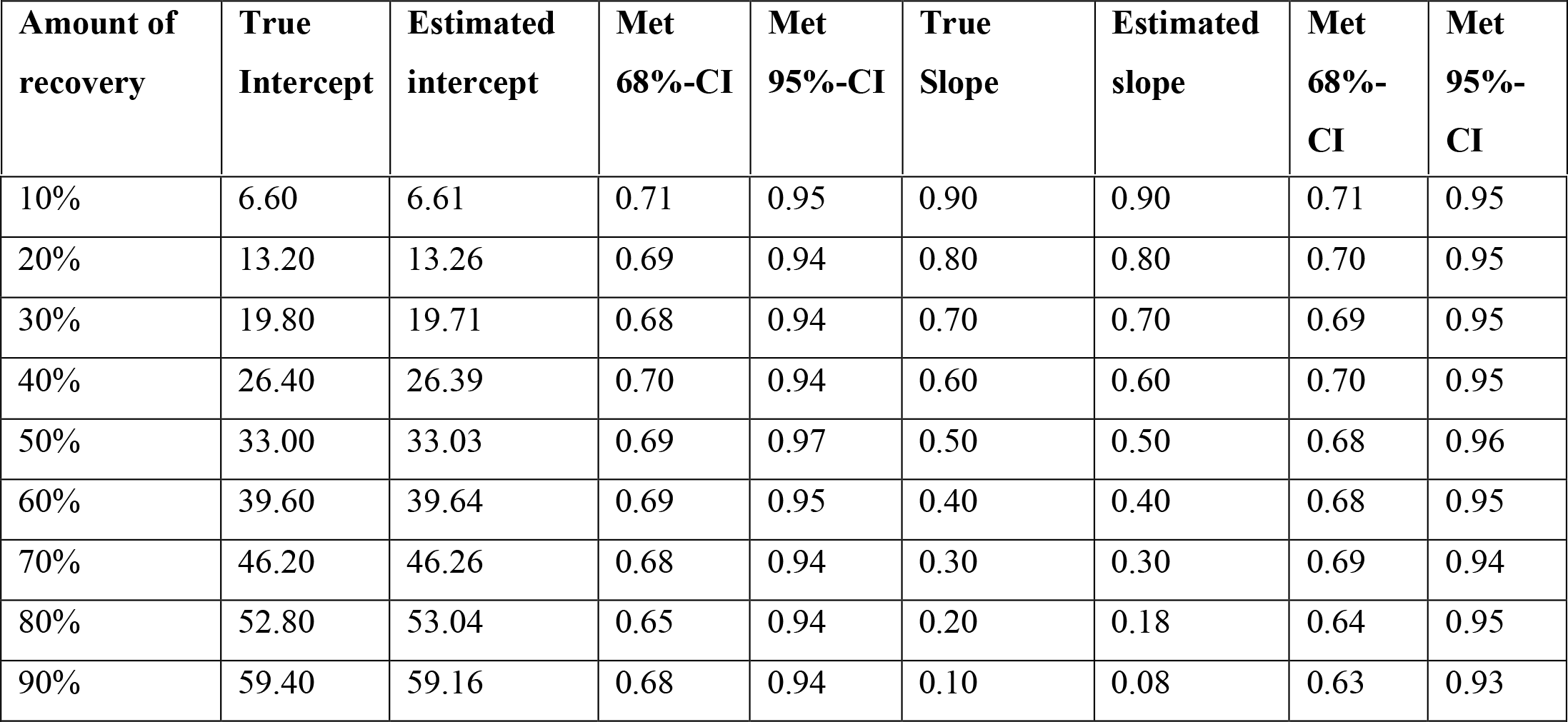
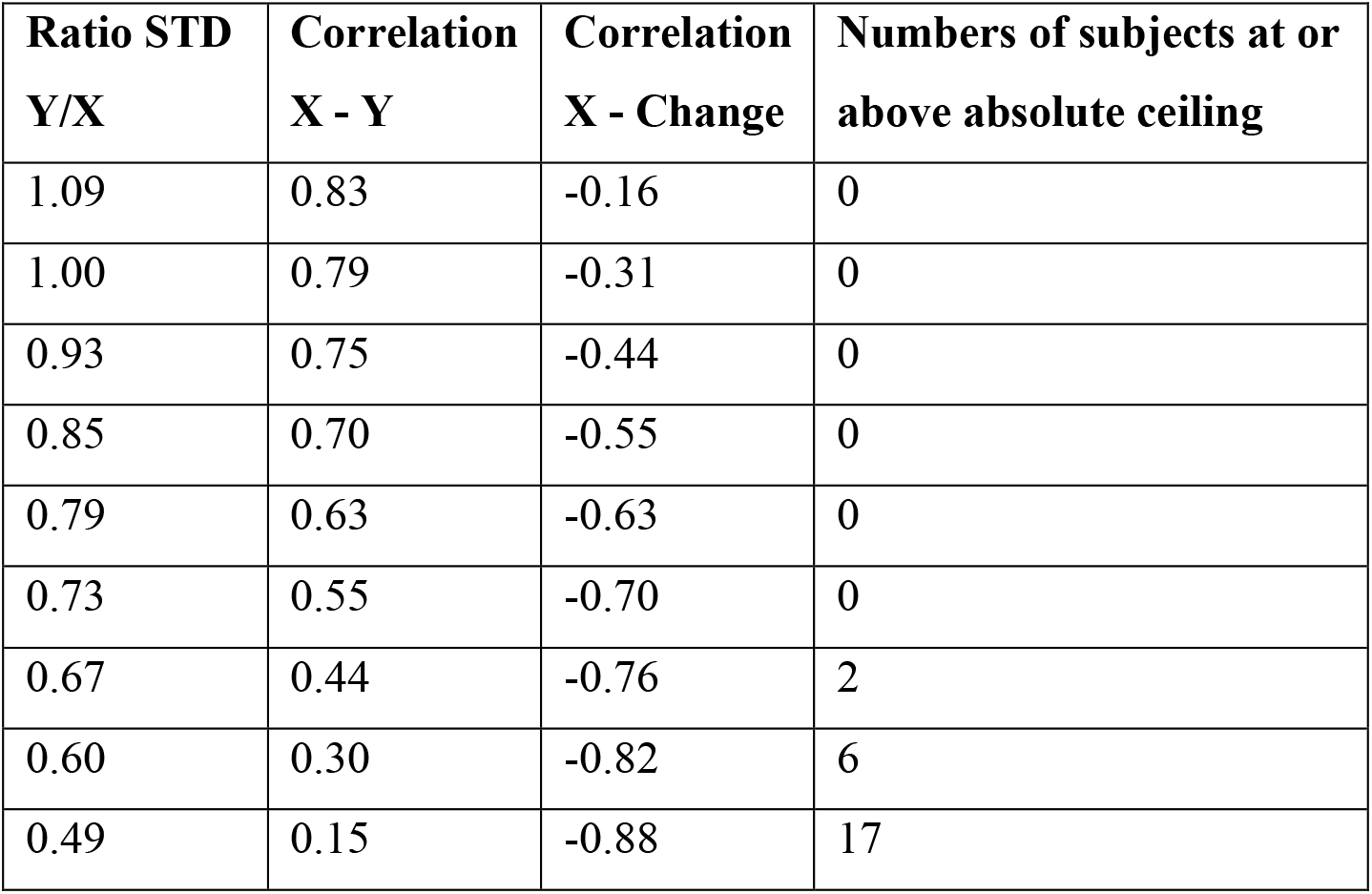
Proportional to lost function recovery: Ceiling enforced, Subset 10 - 40.

| Amount of recovery | True Intercept | Estimated intercept | Met 68%-CI | Met 95%-CI | True Slope | Estimated slope | Met 68%-CI | Met 95%-CI |
| --- | --- | --- | --- | --- | --- | --- | --- | --- |
| 10% | 6.60 | 6.61 | 0.71 | 0.95 | 0.90 | 0.90 | 0.71 | 0.95 |
| 20% | 13.20 | 13.26 | 0.69 | 0.94 | 0.80 | 0.80 | 0.70 | 0.95 |
| 30% | 19.80 | 19.71 | 0.68 | 0.94 | 0.70 | 0.70 | 0.69 | 0.95 |
| 40% | 26.40 | 26.39 | 0.70 | 0.94 | 0.60 | 0.60 | 0.70 | 0.95 |
| 50% | 33.00 | 33.03 | 0.69 | 0.97 | 0.50 | 0.50 | 0.68 | 0.96 |
| 60% | 39.60 | 39.64 | 0.69 | 0.95 | 0.40 | 0.40 | 0.68 | 0.95 |
| 70% | 46.20 | 46.26 | 0.68 | 0.94 | 0.30 | 0.30 | 0.69 | 0.94 |
| 80% | 52.80 | 53.04 | 0.65 | 0.94 | 0.20 | 0.18 | 0.64 | 0.95 |
| 90% | 59.40 | 59.16 | 0.68 | 0.94 | 0.10 | 0.08 | 0.63 | 0.93 |

| **Ratio STD**<br>**Y/X** | **Correlation**<br>**X - Y** | **Correlation**<br>**X - Change** | **Numbers of subjects at or**<br>**above absolute ceiling** |
| --- | --- | --- | --- |
| 1.09 | 0.83 | -0.16 | 0 |
| 1.00 | 0.79 | -0.31 | 0 |
| 0.93 | 0.75 | -0.44 | 0 |
| 0.85 | 0.70 | -0.55 | 0 |
| 0.79 | 0.63 | -0.63 | 0 |
| 0.73 | 0.55 | -0.70 | 0 |
| 0.67 | 0.44 | -0.76 | 2 |
| 0.60 | 0.30 | -0.82 | 6 |
| 0.49 | 0.15 | -0.88 | 17 |

**Supplementary table 1f:** Proportional to lost function recovery: Ceiling enforced, Subset 10 - 45.

| **Amount of**<br>**recovery** | **True**<br>**Intercept** | **Estimated**<br>**intercept** | **Met**<br>**68%-CI** | **Met**<br>**95%-CI** | **True**<br>**Slope** | **Estimated**<br>**slope** | **Met**<br>**68%-**<br>**CI** | **Met**<br>**95%-**<br>**CI** |
| --- | --- | --- | --- | --- | --- | --- | --- | --- |
| 10% | 6.60 | 6.61 | 0.67 | 0.94 | 0.90 | 0.90 | 0.67 | 0.94 |
| 20% | 13.20 | 13.14 | 0.68 | 0.95 | 0.80 | 0.80 | 0.70 | 0.95 |
| 30% | 19.80 | 19.77 | 0.69 | 0.95 | 0.70 | 0.70 | 0.68 | 0.95 |
| 40% | 26.40 | 26.49 | 0.68 | 0.95 | 0.60 | 0.60 | 0.67 | 0.95 |
| 50% | 33.00 | 33.08 | 0.71 | 0.96 | 0.50 | 0.50 | 0.71 | 0.96 |
| 60% | 39.60 | 39.66 | 0.69 | 0.95 | 0.40 | 0.40 | 0.70 | 0.95 |
| 70% | 46.20 | 46.38 | 0.66 | 0.94 | 0.30 | 0.29 | 0.64 | 0.94 |
| 80% | 52.80 | 53.03 | 0.68 | 0.95 | 0.20 | 0.18 | 0.66 | 0.93 |
| 90% | 59.40 | 59.25 | 0.68 | 0.94 | 0.10 | 0.07 | 0.54 | 0.89 |

| Ratio STD<br>Y/X | Correlation<br>X - Y | Correlation<br>X - Change | Numbers of subjects at or<br>above absolute ceiling |
| --- | --- | --- | --- |
| 1.03 | 0.87 | -0.19 | 0 |
| 0.95 | 0.84 | -0.36 | 0 |
| 0.87 | 0.81 | -0.50 | 0 |
| 0.79 | 0.76 | -0.62 | 0 |
| 0.71 | 0.70 | -0.70 | 0 |
| 0.65 | 0.61 | -0.76 | 1 |
| 0.58 | 0.50 | -0.82 | 3 |
| 0.51 | 0.35 | -0.87 | 8 |
| 0.41 | 0.17 | -0.92 | 20 |

### Proportional to spared function recovery

#### Baseline models (state noise only)

It is important to note that – in contrast to proportional to lost function – when formulated in standard-form, proportional to spared function is characterized by intercepts of zero and slopes greater than one (and less than 2); see **Figure 2, panel [E]**. Pearson correlations of *FM-initial – Change* as well as *FM-end – FM-initial* reached maximal values of almost 1 for increasing degrees of proportional to spared function recovery. These were accompanied by ratios of standard deviations greater than one (90 %: 1.9) (c.f. **Figure 6, B and C**, center plots). This is the regime “on the hill” in our surface plot, i.e., log variability ratio substantially greater than zero, **Figure 1**.

#### Ceiling enforced

Setting any *FM-end* values greater than it to 66 had a detrimental effect on the accurate estimation of parameters for all proportional to spared instantiations, confidence intervals were not met in any of the cases. Crucially, estimated intercept and slope combinations resembled those of the true constant and proportional to lost function recovery models: Intercepts increased in parallel to increasing amounts of proportional to spared function, while slopes started at values close to one and successively decreased to a minimum of 0.76. Even in the case of only 10 % proportional to spared function, there were 9.1 % of subjects at absolute ceiling (maximum: 90 % proportional to spared function recovery with 30.9 % subjects at ceiling).

#### Subset approaches FM-initial 10 to 60, 50, 40

Once again, accuracy of estimation performance increased with decreasing upper bounds for the FM-initial scores, yet not as completely as for proportional to lost function: *FM-initial* 10 – 60: up to 10 %, *FM-initial* 10 – 50: up to 30 % and *FM-initial* 10 – 40: up to 70 % of proportional to spared function recovery.

**Supplementary table 2a:** Proportional to spared function recovery: No ceiling enforced.

| Amount of recovery | True Intercept | Estimated intercept | Met 68%-CI | Met 95%-CI | True Slope | Estimated slope | Met 68%-CI | Met 95%-CI |
| --- | --- | --- | --- | --- | --- | --- | --- | --- |
| 10% | 0.00 | 0.00 | 0.67 | 0.94 | 1.10 | 1.10 | 0.71 | 0.94 |
| 20% | 0.00 | 0.01 | 0.70 | 0.95 | 1.20 | 1.20 | 0.68 | 0.95 |
| 30% | 0.00 | 0.01 | 0.71 | 0.95 | 1.30 | 1.30 | 0.71 | 0.95 |
| 40% | 0.00 | -0.01 | 0.67 | 0.95 | 1.40 | 1.40 | 0.68 | 0.96 |
| 50% | 0.00 | 0.00 | 0.67 | 0.95 | 1.50 | 1.50 | 0.67 | 0.94 |
| 60% | 0.00 | 0.00 | 0.69 | 0.95 | 1.60 | 1.60 | 0.69 | 0.95 |
| 70% | 0.00 | -0.02 | 0.71 | 0.96 | 1.70 | 1.70 | 0.68 | 0.96 |
| 80% | 0.00 | -0.02 | 0.67 | 0.94 | 1.80 | 1.80 | 0.68 | 0.94 |
| 90% | 0.00 | 0.00 | 0.70 | 0.96 | 1.90 | 1.90 | 0.68 | 0.96 |

| Ratio STD Y/X | Correlation X - Y | Correlation X - Change |
| --- | --- | --- |
| 1.11 | 0.99 | 0.66 |
| 1.21 | 1.00 | 0.87 |
| 1.30 | 1.00 | 0.93 |
| 1.40 | 1.00 | 0.96 |
| 1.50 | 1.00 | 0.98 |
| 1.60 | 1.00 | 0.98 |
| 1.70 | 1.00 | 0.99 |
| 1.80 | 1.00 | 0.99 |
| 1.90 | 1.00 | 0.99 |

**Supplementary table 2b:**
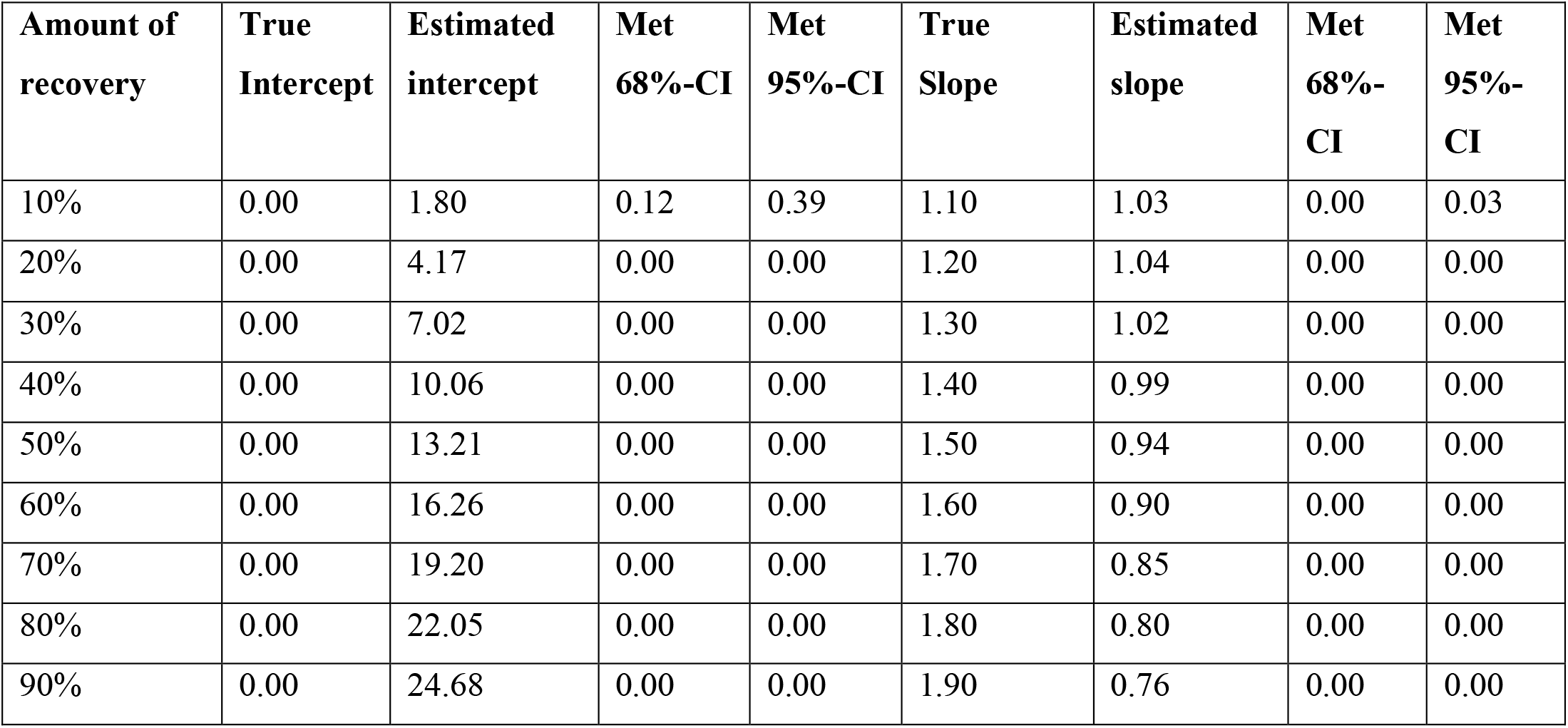
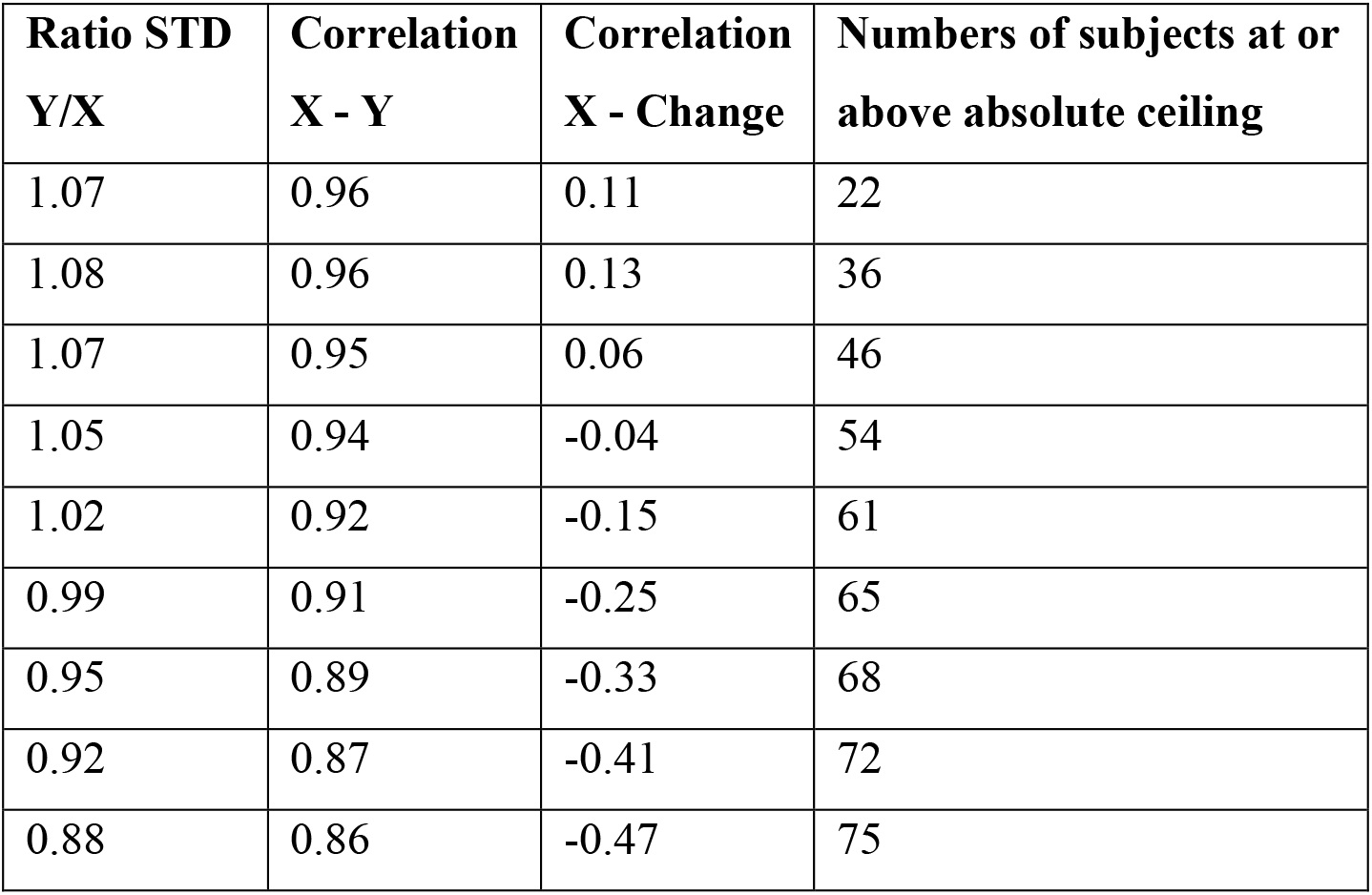
Proportional to spared function recovery: Ceiling enforced.

**Supplementary table 2c:** Proportional to spared function recovery: Ceiling enforced, Subset 10 - 60.

| Amount of recovery | True Intercept | Estimated intercept | Met 68%-CI | Met 95%-CI | True Slope | Estimated slope | Met 68%-CI | Met 95%-CI |
| --- | --- | --- | --- | --- | --- | --- | --- | --- |
| 10% | 0.00 | 0.46 | 0.61 | 0.92 | 1.10 | 1.08 | 0.54 | 0.87 |
| 20% | 0.00 | 1.73 | 0.17 | 0.50 | 1.20 | 1.13 | 0.00 | 0.08 |
| 30% | 0.00 | 3.75 | 0.00 | 0.01 | 1.30 | 1.14 | 0.00 | 0.00 |
| 40% | 0.00 | 6.42 | 0.00 | 0.00 | 1.40 | 1.12 | 0.00 | 0.00 |
| 50% | 0.00 | 9.30 | 0.00 | 0.00 | 1.50 | 1.09 | 0.00 | 0.00 |
| 60% | 0.00 | 12.26 | 0.00 | 0.00 | 1.60 | 1.04 | 0.00 | 0.00 |
| 70% | 0.00 | 15.24 | 0.00 | 0.00 | 1.70 | 1.00 | 0.00 | 0.00 |
| 80% | 0.00 | 18.09 | 0.00 | 0.00 | 1.80 | 0.95 | 0.00 | 0.00 |
| 90% | 0.00 | 20.82 | 0.00 | 0.00 | 1.90 | 0.90 | 0.00 | 0.00 |

**Supplementary table 2d:** Proportional to spared function recovery: Ceiling enforced, Subset 10 - 50.

| **Amount of**<br>**recovery** | **True**<br>**Intercept** | **Estimated**<br>**intercept** | **Met**<br>**68%-CI** | **Met**<br>**95%-CI** | **True**<br>**Slope** | **Estimated**<br>**slope** | **Met**<br>**68%-**<br>**CI** | **Met**<br>**95%-**<br>**CI** |
| --- | --- | --- | --- | --- | --- | --- | --- | --- |
| 10% | 0.00 | 0.04 | 0.69 | 0.95 | 1.10 | 1.10 | 0.66 | 0.95 |
| 20% | 0.00 | 0.14 | 0.67 | 0.95 | 1.20 | 1.19 | 0.68 | 0.94 |
| 30% | 0.00 | 0.38 | 0.69 | 0.93 | 1.30 | 1.28 | 0.64 | 0.92 |
| 40% | 0.00 | 1.31 | 0.37 | 0.75 | 1.40 | 1.34 | 0.15 | 0.51 |
| 50% | 0.00 | 3.08 | 0.03 | 0.18 | 1.50 | 1.35 | 0.00 | 0.00 |
| 60% | 0.00 | 5.24 | 0.00 | 0.00 | 1.60 | 1.34 | 0.00 | 0.00 |
| 70% | 0.00 | 7.72 | 0.00 | 0.00 | 1.70 | 1.31 | 0.00 | 0.00 |
| 80% | 0.00 | 10.40 | 0.00 | 0.00 | 1.80 | 1.28 | 0.00 | 0.00 |
| 90% | 0.00 | 13.02 | 0.00 | 0.00 | 1.90 | 1.23 | 0.00 | 0.00 |

| **Ratio STD**<br>**Y/X** | **Correlation**<br>**X - Y** | **Correlation**<br>**X - Change** | **Numbers of subjects at or**<br>**above absolute ceiling** |
| --- | --- | --- | --- |
| 1.18 | 0.93 | 0.22 | 0 |
| 1.27 | 0.94 | 0.42 | 1 |
| 1.35 | 0.95 | 0.56 | 5 |
| 1.39 | 0.96 | 0.64 | 10 |
| 1.41 | 0.96 | 0.66 | 16 |
| 1.41 | 0.95 | 0.62 | 20 |
| 1.39 | 0.94 | 0.57 | 23 |
| 1.37 | 0.93 | 0.49 | 27 |
| 1.34 | 0.92 | 0.41 | 30 |

**Supplementary table 2e:** Proportional to spared function recovery: Ceiling enforced, Subset 10 - 40.

| **Amount of**<br>**recovery** | **True**<br>**Intercept** | **Estimated**<br>**intercept** | **Met**<br>**68%-CI** | **Met**<br>**95%-CI** | **True**<br>**Slope** | **Estimated**<br>**slope** | **Met**<br>**68%-**<br>**CI** | **Met**<br>**95%-**<br>**CI** |
| --- | --- | --- | --- | --- | --- | --- | --- | --- |
| 10% | 0.00 | -0.02 | 0.68 | 0.96 | 1.10 | 1.10 | 0.69 | 0.95 |
| 20% | 0.00 | -0.01 | 0.66 | 0.94 | 1.20 | 1.20 | 0.68 | 0.94 |
| 30% | 0.00 | -0.02 | 0.71 | 0.96 | 1.30 | 1.30 | 0.72 | 0.95 |
| 40% | 0.00 | -0.04 | 0.67 | 0.95 | 1.40 | 1.40 | 0.67 | 0.95 |
| 50% | 0.00 | 0.04 | 0.70 | 0.94 | 1.50 | 1.50 | 0.68 | 0.95 |
| 60% | 0.00 | 0.20 | 0.67 | 0.95 | 1.60 | 1.59 | 0.67 | 0.95 |
| 70% | 0.00 | 0.52 | 0.64 | 0.93 | 1.70 | 1.67 | 0.61 | 0.92 |
| 80% | 0.00 | 1.39 | 0.47 | 0.83 | 1.80 | 1.72 | 0.33 | 0.71 |
| 90% | 0.00 | 2.78 | 0.14 | 0.47 | 1.90 | 1.74 | 0.01 | 0.12 |

| **Ratio STD**<br>**Y/X** | **Correlation**<br>**X - Y** | **Correlation**<br>**X - Change** | **Numbers of subjects at or**<br>**above absolute ceiling** |
| --- | --- | --- | --- |
| 1.26 | 0.87 | 0.16 | 0 |
| 1.34 | 0.89 | 0.31 | 0 |
| 1.44 | 0.91 | 0.44 | 0 |
| 1.53 | 0.92 | 0.55 | 0 |
| 1.62 | 0.93 | 0.63 | 0 |
| 1.70 | 0.94 | 0.70 | 1 |
| 1.77 | 0.94 | 0.75 | 3 |
| 1.82 | 0.95 | 0.78 | 7 |
| 1.83 | 0.95 | 0.79 | 10 |

**Supplementary table 2f:** Proportional to spared function recovery: Ceiling enforced, Subset 10 - 45.

| Amount of recovery | True Intercept | Estimated intercept | Met 68%-CI | Met 95%-CI | True Slope | Estimated slope | Met 68%-CI | Met 95%-CI |
| --- | --- | --- | --- | --- | --- | --- | --- | --- |
| 10% | 0.00 | 0.04 | 0.67 | 0.93 | 1.10 | 1.10 | 0.68 | 0.93 |
| 20% | 0.00 | -0.02 | 0.70 | 0.95 | 1.20 | 1.20 | 0.69 | 0.95 |
| 30% | 0.00 | 0.03 | 0.67 | 0.94 | 1.30 | 1.30 | 0.69 | 0.94 |
| 40% | 0.00 | 0.19 | 0.70 | 0.94 | 1.40 | 1.39 | 0.69 | 0.95 |
| 50% | 0.00 | 0.74 | 0.59 | 0.90 | 1.50 | 1.46 | 0.56 | 0.87 |
| 60% | 0.00 | 1.79 | 0.30 | 0.70 | 1.60 | 1.51 | 0.09 | 0.44 |
| 70% | 0.00 | 3.32 | 0.04 | 0.24 | 1.70 | 1.53 | 0.00 | 0.01 |
| 80% | 0.00 | 5.25 | 0.00 | 0.01 | 1.80 | 1.52 | 0.00 | 0.00 |
| 90% | 0.00 | 7.47 | 0.00 | 0.00 | 1.90 | 1.50 | 0.00 | 0.00 |

| Ratio STD Y/X | Correlation X - Y | Correlation X - Change | Numbers of subjects at or above absolute ceiling |
| --- | --- | --- | --- |
| 1.21 | 0.91 | 0.19 | 0 |
| 1.31 | 0.92 | 0.36 | 0 |
| 1.39 | 0.93 | 0.50 | 0 |
| 1.48 | 0.94 | 0.61 | 2 |
| 1.54 | 0.95 | 0.68 | 4 |
| 1.59 | 0.95 | 0.71 | 7 |
| 1.61 | 0.95 | 0.72 | 10 |
| 1.61 | 0.94 | 0.70 | 14 |
| 1.60 | 0.94 | 0.67 | 17 |

### Constant recovery

#### Baseline models (state noise only)

When formulated in standard-form regression, the natural behavior of constant models is expressed by increasing intercepts, with unvarying slopes at one; see **Figure 2[F]**. Pearson correlations for *FM-end – FM-initial*, as well as the ratio of standard deviations, occupy values close to 1, *FM-initial – Change*, in contrast, are not correlated; these phenomena are independent of the amount of constant recovery (c.f. **Figure 6, B and C**, right plots).

#### Ceiling enforced

The ceiling effect rendered it impossible to veridically estimate the true model parameters. Confidence intervals were not met in any simulation run for any constant model above 5 points recovery. Of further note, the ratio of standard deviations decreased from 0.99 to a minimum of 0.13; see **Figure 6 [C]**, rightmost panel, thus becoming even more extreme than the ratio for proportional to lost function. A dissociation of correlations also reflected this: the correlation of *FM-initial – Change* dropped from zero to an almost perfect anticorrelation of -1, while the correlation of *FM-end – FM-initial* showed the opposite behavior, starting at 0.96, yet dropping to 0.36 for a maximum of 50 points constant recovery; see **Figure 6 [B]** rightmost panel. The percentages of subjects at ceiling increased from 8.2 % to 45.3 %.

#### Subset approaches FM-initial 10 to 60, 50, 40

Reducing analyses to subjects with initial FM scores of 10 – 60 only marginally improved parameter estimation performance, i.e., up to 5 points constant recovery. Considering subjects with initial FM scores of 10 – 50 and 10 – 40 allowed accurate estimation up to 10 – 15 points and 25 points, respectively. Noteworthily, estimation performance dropped once the percentage of subjects at ceiling exceeded 7 %.

Exclusively looking at subjects in the range of *FM-initial* 10 – 60 reduced the empirical number of subjects at absolute ceiling from 37 to 18 (15.2 % and 9.0 %, respectively), in the range of *FM-initial* 10 – 50 to 7 (4.7 %) and in the range of *FM-initial* 10 – 40 to 4 (4.3 %).

**Supplementary table 3a:** Constant recovery: No ceiling enforced.

| Amount of recovery | True Intercept | Estimated intercept | Met 68%-CI | Met 95%-CI | True Slope | Estimated slope | Met 68%-CI | Met 95%-CI |
| --- | --- | --- | --- | --- | --- | --- | --- | --- |
| 5 | 5.00 | 4.99 | 0.70 | 0.95 | 1.00 | 1.00 | 0.69 | 0.95 |
| 10 | 10.00 | 10.00 | 0.67 | 0.94 | 1.00 | 1.00 | 0.68 | 0.95 |
| 15 | 15.00 | 14.97 | 0.70 | 0.94 | 1.00 | 1.00 | 0.68 | 0.95 |
| 20 | 20.00 | 19.99 | 0.68 | 0.94 | 1.00 | 1.00 | 0.67 | 0.94 |
| 25 | 25.00 | 24.99 | 0.71 | 0.95 | 1.00 | 1.00 | 0.70 | 0.95 |
| 30 | 30.00 | 29.98 | 0.69 | 0.95 | 1.00 | 1.00 | 0.67 | 0.95 |
| 35 | 35.00 | 34.99 | 0.69 | 0.95 | 1.00 | 1.00 | 0.68 | 0.95 |
| 40 | 40.00 | 40.01 | 0.69 | 0.96 | 1.00 | 1.00 | 0.66 | 0.95 |
| 45 | 45.00 | 45.03 | 0.70 | 0.95 | 1.00 | 1.00 | 0.68 | 0.95 |
| 50 | 50.00 | 50.01 | 0.69 | 0.95 | 1.00 | 1.00 | 0.68 | 0.95 |

| Ratio STD Y/X | Correlation X - Y | Correlation X - Change |
| --- | --- | --- |
| 1.04 | 0.96 | 0.00 |
| 1.04 | 0.96 | 0.00 |
| 1.04 | 0.96 | 0.00 |
| 1.04 | 0.96 | 0.00 |
| 1.04 | 0.96 | 0.00 |
| 1.04 | 0.96 | 0.00 |
| 1.04 | 0.96 | 0.00 |
| 1.04 | 0.96 | 0.00 |
| 1.04 | 0.96 | 0.00 |
| 1.04 | 0.96 | 0.00 |

**Supplementary table 3b:**
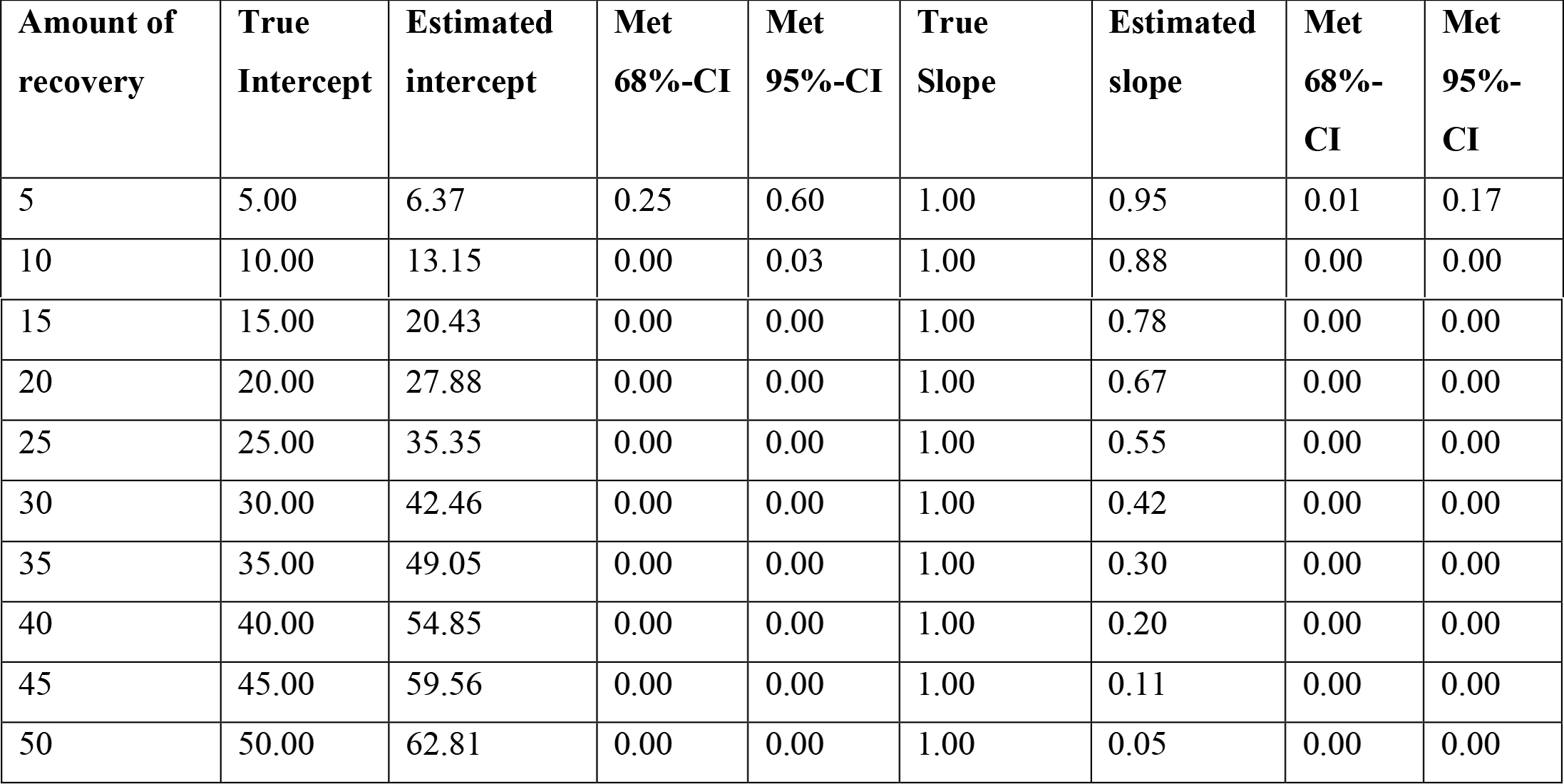
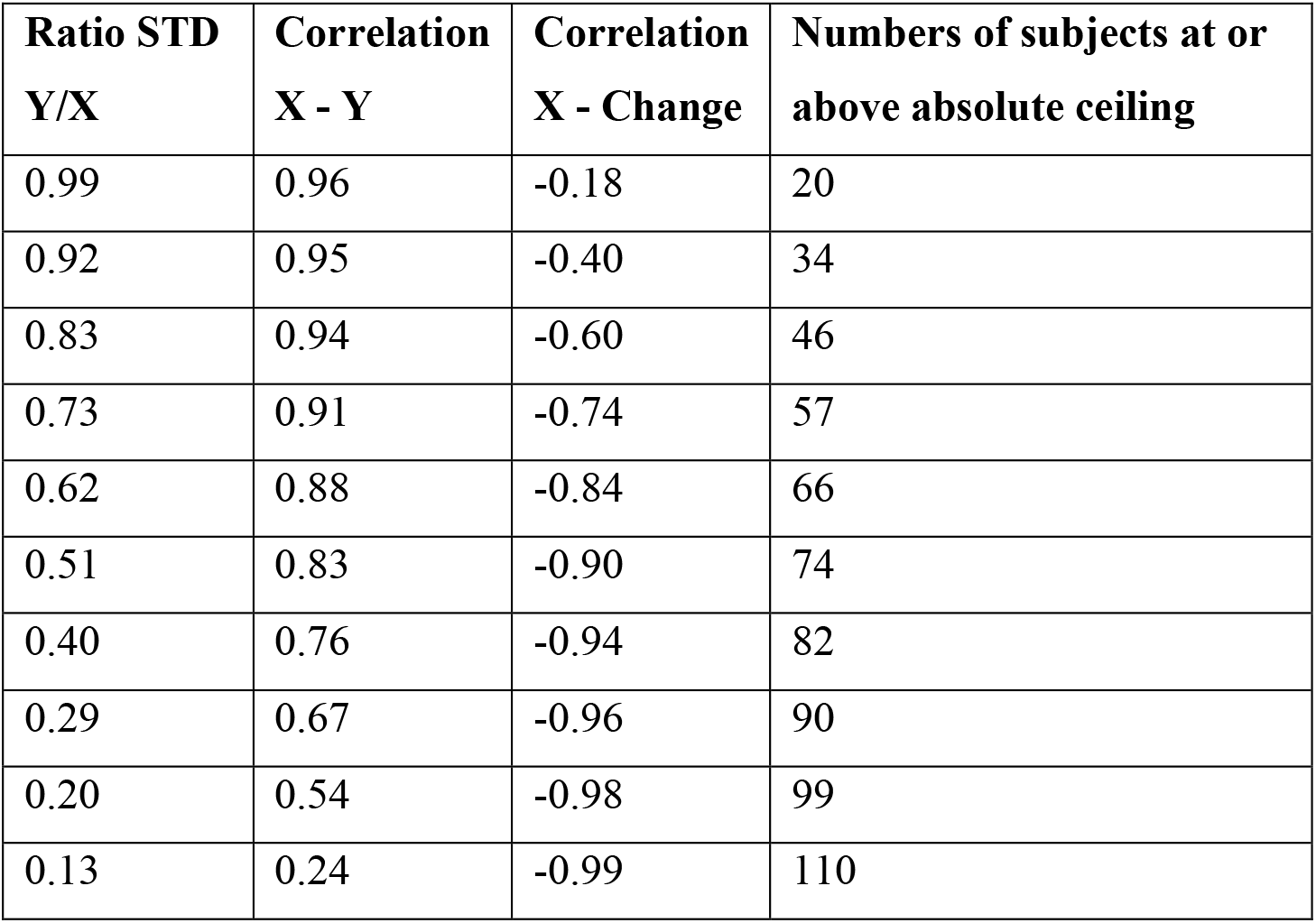
Constant recovery: Ceiling enforced.

| Amount of recovery | True Intercept | Estimated intercept | Met 68%-CI | Met 95%-CI | True Slope | Estimated slope | Met 68%-CI | Met 95%-CI |
| --- | --- | --- | --- | --- | --- | --- | --- | --- |
| 5 | 5.00 | 6.37 | 0.25 | 0.60 | 1.00 | 0.95 | 0.01 | 0.17 |
| 10 | 10.00 | 13.15 | 0.00 | 0.03 | 1.00 | 0.88 | 0.00 | 0.00 |
| 15 | 15.00 | 20.43 | 0.00 | 0.00 | 1.00 | 0.78 | 0.00 | 0.00 |
| 20 | 20.00 | 27.88 | 0.00 | 0.00 | 1.00 | 0.67 | 0.00 | 0.00 |
| 25 | 25.00 | 35.35 | 0.00 | 0.00 | 1.00 | 0.55 | 0.00 | 0.00 |
| 30 | 30.00 | 42.46 | 0.00 | 0.00 | 1.00 | 0.42 | 0.00 | 0.00 |
| 35 | 35.00 | 49.05 | 0.00 | 0.00 | 1.00 | 0.30 | 0.00 | 0.00 |
| 40 | 40.00 | 54.85 | 0.00 | 0.00 | 1.00 | 0.20 | 0.00 | 0.00 |
| 45 | 45.00 | 59.56 | 0.00 | 0.00 | 1.00 | 0.11 | 0.00 | 0.00 |
| 50 | 50.00 | 62.81 | 0.00 | 0.00 | 1.00 | 0.05 | 0.00 | 0.00 |

| **Ratio STD**<br>**Y/X** | **Correlation**<br>**X - Y** | **Correlation**<br>**X - Change** | **Numbers of subjects at or**<br>**above absolute ceiling** |
| --- | --- | --- | --- |
| 0.99 | 0.96 | -0.18 | 20 |
| 0.92 | 0.95 | -0.40 | 34 |
| 0.83 | 0.94 | -0.60 | 46 |
| 0.73 | 0.91 | -0.74 | 57 |
| 0.62 | 0.88 | -0.84 | 66 |
| 0.51 | 0.83 | -0.90 | 74 |
| 0.40 | 0.76 | -0.94 | 82 |
| 0.29 | 0.67 | -0.96 | 90 |
| 0.20 | 0.54 | -0.98 | 99 |
| 0.13 | 0.24 | -0.99 | 110 |

**Supplementary table 3c:**
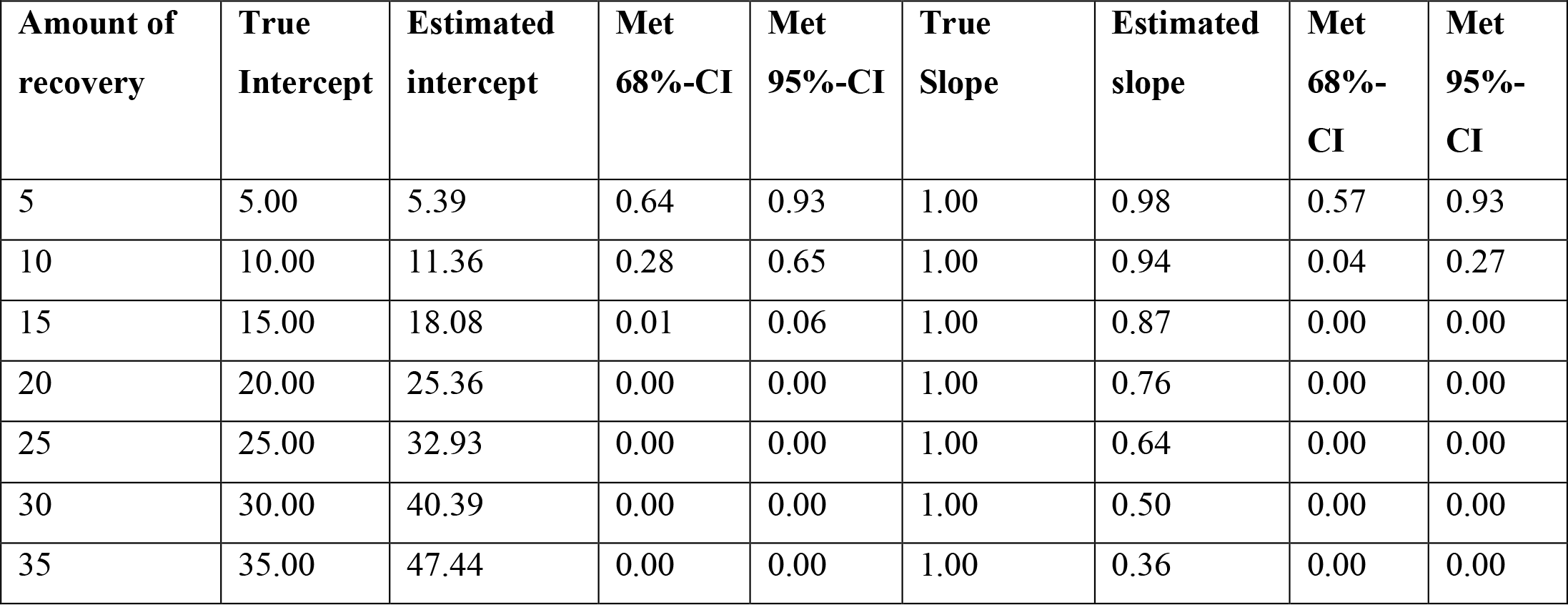
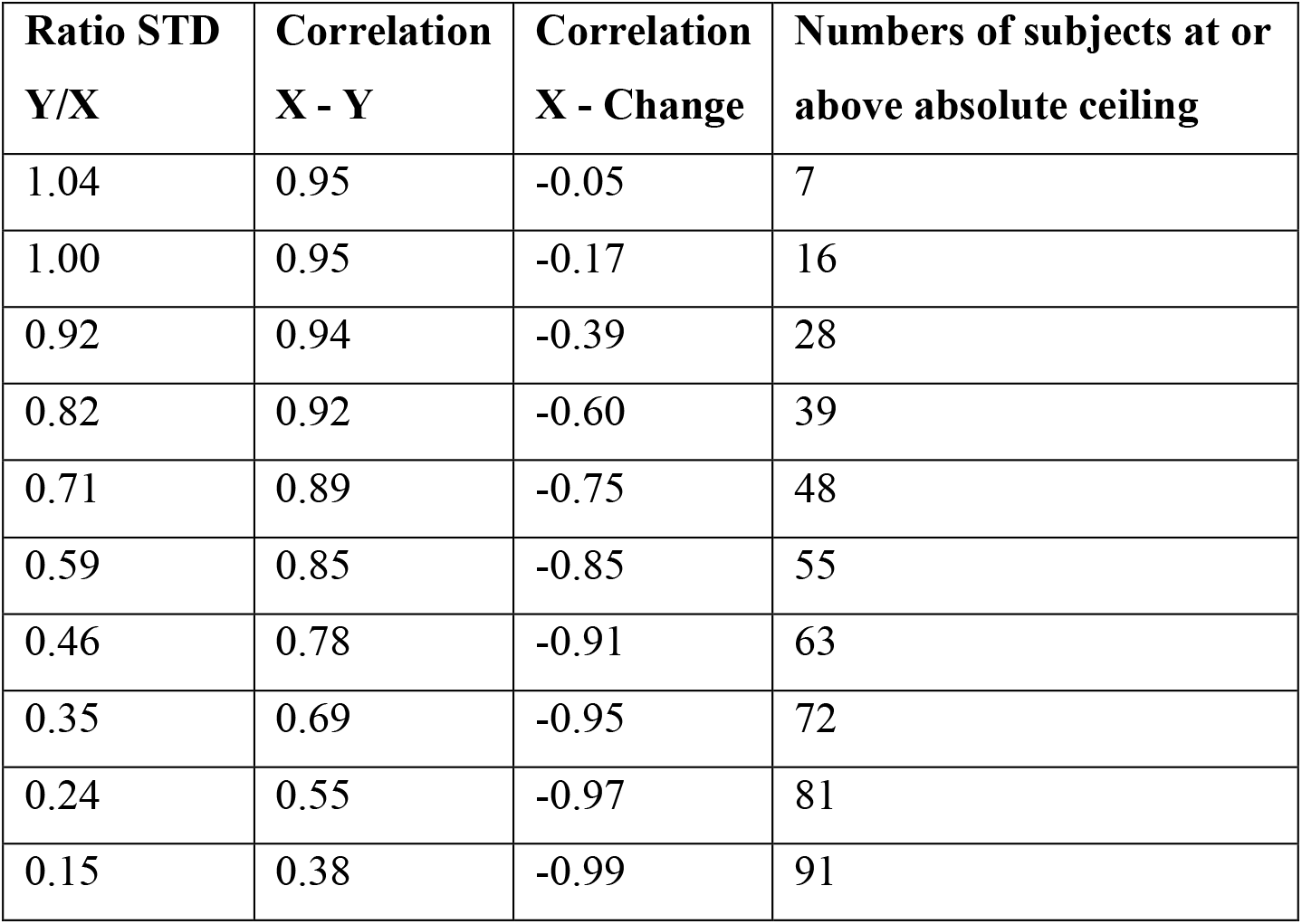
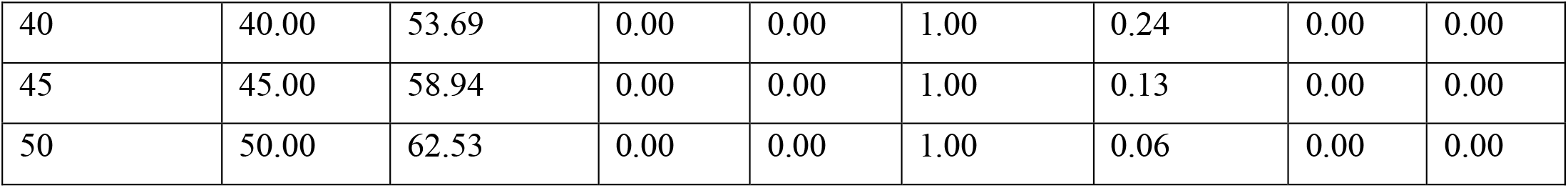
Constant recovery: Ceiling enforced, Subset 10 - 60.

**Supplementary table 3d:** Constant recovery: Ceiling enforced, Subset 10 - 50.

| **Amount of**<br>**recovery** | **True**<br>**Intercept** | **Estimated**<br>**intercept** | **Met**<br>**68%-CI** | **Met**<br>**95%-CI** | **True**<br>**Slope** | **Estimated**<br>**slope** | **Met**<br>**68%-**<br>**CI** | **Met**<br>**95%-**<br>**CI** |
| --- | --- | --- | --- | --- | --- | --- | --- | --- |
| 5 | 5.00 | 5.02 | 0.67 | 0.96 | 1.00 | 1.00 | 0.69 | 0.95 |
| 10 | 10.00 | 10.12 | 0.68 | 0.95 | 1.00 | 0.99 | 0.69 | 0.96 |
| 15 | 15.00 | 15.46 | 0.66 | 0.94 | 1.00 | 0.98 | 0.61 | 0.91 |
| 20 | 20.00 | 21.63 | 0.28 | 0.64 | 1.00 | 0.92 | 0.05 | 0.26 |
| 25 | 25.00 | 28.71 | 0.00 | 0.05 | 1.00 | 0.81 | 0.00 | 0.00 |
| 30 | 30.00 | 36.35 | 0.00 | 0.00 | 1.00 | 0.67 | 0.00 | 0.00 |
| 35 | 35.00 | 44.00 | 0.00 | 0.00 | 1.00 | 0.51 | 0.00 | 0.00 |
| 40 | 40.00 | 51.23 | 0.00 | 0.00 | 1.00 | 0.34 | 0.00 | 0.00 |
| 45 | 45.00 | 57.42 | 0.00 | 0.00 | 1.00 | 0.20 | 0.00 | 0.00 |
| **50** | 50.00 | 61.87 | 0.00 | 0.00 | 1.00 | 0.09 | 0.00 | 0.00 |

| **Ratio STD**<br>**Y/X** | **Correlation**<br>**X - Y** | **Correlation**<br>**X - Change** | **Numbers of subjects at or**<br>**above absolute ceiling** |
| --- | --- | --- | --- |
| 1.09 | 0.92 | 0.00 | 0 |
| 1.08 | 0.92 | -0.01 | 1 |
| 1.06 | 0.92 | -0.05 | 6 |
| 1.00 | 0.92 | -0.20 | 13 |
| 0.90 | 0.90 | -0.43 | 21 |
| 0.77 | 0.87 | -0.65 | 29 |
| 0.63 | 0.81 | -0.80 | 37 |
| 0.48 | 0.72 | -0.89 | 45 |
| 0.33 | 0.58 | -0.95 | 55 |
| 0.21 | 0.40 | -0.98 | 64 |

**Supplementary table 3e:**
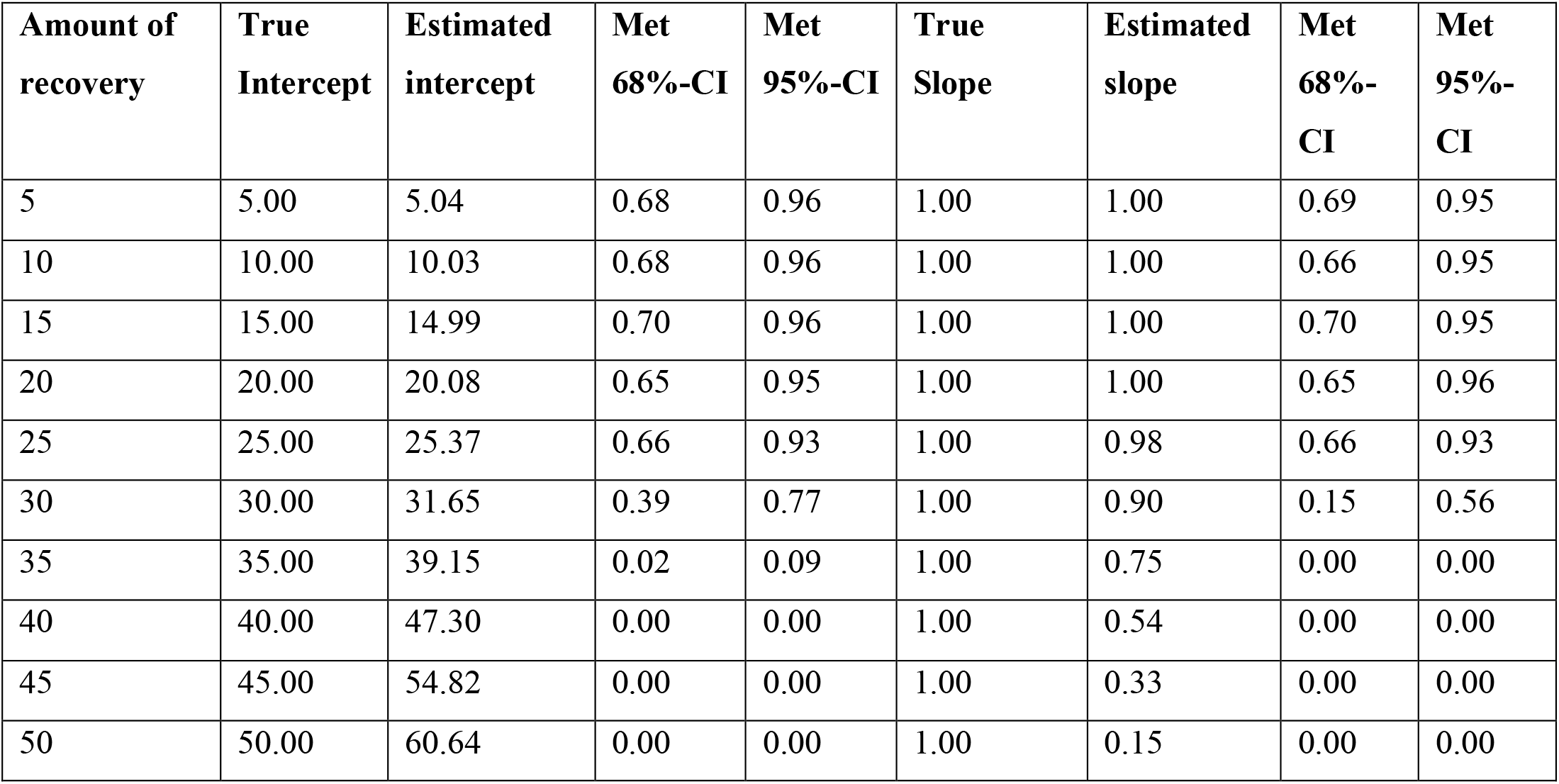
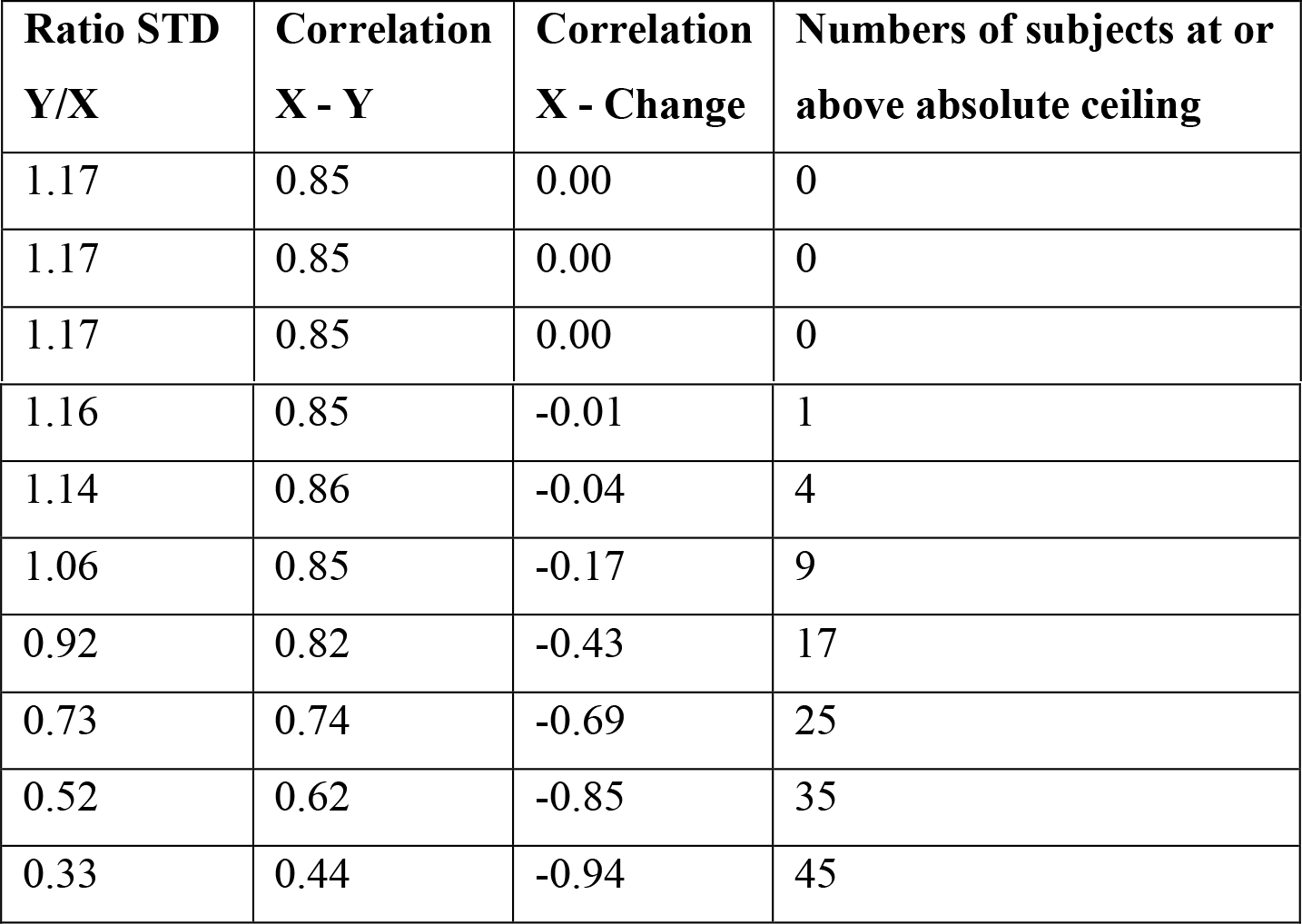
Constant recovery: Ceiling enforced, Subset 10 - 40.

| **Amount of**<br>**recovery** | **True**<br>**Intercept** | **Estimated**<br>**intercept** | **Met**<br>**68%-CI** | **Met**<br>**95%-CI** | **True**<br>**Slope** | **Estimated**<br>**slope** | **Met**<br>**68%-**<br>**CI** | **Met**<br>**95%-**<br>**CI** |
| --- | --- | --- | --- | --- | --- | --- | --- | --- |
| 5 | 5.00 | 5.04 | 0.68 | 0.96 | 1.00 | 1.00 | 0.69 | 0.95 |
| 10 | 10.00 | 10.03 | 0.68 | 0.96 | 1.00 | 1.00 | 0.66 | 0.95 |
| 15 | 15.00 | 14.99 | 0.70 | 0.96 | 1.00 | 1.00 | 0.70 | 0.95 |
| 20 | 20.00 | 20.08 | 0.65 | 0.95 | 1.00 | 1.00 | 0.65 | 0.96 |
| 25 | 25.00 | 25.37 | 0.66 | 0.93 | 1.00 | 0.98 | 0.66 | 0.93 |
| 30 | 30.00 | 31.65 | 0.39 | 0.77 | 1.00 | 0.90 | 0.15 | 0.56 |
| 35 | 35.00 | 39.15 | 0.02 | 0.09 | 1.00 | 0.75 | 0.00 | 0.00 |
| 40 | 40.00 | 47.30 | 0.00 | 0.00 | 1.00 | 0.54 | 0.00 | 0.00 |
| 45 | 45.00 | 54.82 | 0.00 | 0.00 | 1.00 | 0.33 | 0.00 | 0.00 |
| 50 | 50.00 | 60.64 | 0.00 | 0.00 | 1.00 | 0.15 | 0.00 | 0.00 |

| **Ratio STD**<br>**Y/X** | **Correlation**<br>**X - Y** | **Correlation**<br>**X - Change** | **Numbers of subjects at or**<br>**above absolute ceiling** |
| --- | --- | --- | --- |
| 1.17 | 0.85 | 0.00 | 0 |
| 1.17 | 0.85 | 0.00 | 0 |
| 1.17 | 0.85 | 0.00 | 0 |
| 1.16 | 0.85 | -0.01 | 1 |
| 1.14 | 0.86 | -0.04 | 4 |
| 1.06 | 0.85 | -0.17 | 9 |
| 0.92 | 0.82 | -0.43 | 17 |
| 0.73 | 0.74 | -0.69 | 25 |
| 0.52 | 0.62 | -0.85 | 35 |
| 0.33 | 0.44 | -0.94 | 45 |

**Supplementary table 3f:** Constant recovery: Ceiling enforced, Subset 10 - 45.

| **Amount of recovery** | **True Intercept** | **Estimated intercept** | **Met 68%-CI** | **Met 95%-CI** | **True Slope** | **Estimated slope** | **Met 68%-CI** | **Met 95%-CI** |
| --- | --- | --- | --- | --- | --- | --- | --- | --- |
| 5 | 5.00 | 5.01 | 0.68 | 0.94 | 1.00 | 1.00 | 0.67 | 0.94 |
| 10 | 10.00 | 10.03 | 0.69 | 0.96 | 1.00 | 1.00 | 0.70 | 0.96 |
| 15 | 15.00 | 15.05 | 0.70 | 0.95 | 1.00 | 1.00 | 0.68 | 0.94 |
| 20 | 20.00 | 20.51 | 0.63 | 0.93 | 1.00 | 0.97 | 0.63 | 0.92 |
| 25 | 25.00 | 26.69 | 0.32 | 0.71 | 1.00 | 0.91 | 0.10 | 0.45 |
| 30 | 30.00 | 33.75 | 0.01 | 0.10 | 1.00 | 0.80 | 0.00 | 0.00 |
| 35 | 35.00 | 41.51 | 0.00 | 0.00 | 1.00 | 0.63 | 0.00 | 0.00 |
| 40 | 40.00 | 49.31 | 0.00 | 0.00 | 1.00 | 0.44 | 0.00 | 0.00 |
| 45 | 45.00 | 56.14 | 0.00 | 0.00 | 1.00 | 0.26 | 0.00 | 0.00 |
| 50 | 50.00 | 61.26 | 0.00 | 0.00 | 1.00 | 0.11 | 0.00 | 0.00 |

| **Ratio STD Y/X** | **Correlation X - Y** | **Correlation X - Change** | **Numbers of subjects at or above absolute ceiling** |
| --- | --- | --- | --- |
| 1.12 | 0.89 | 0.00 | 0 |
| 1.12 | 0.89 | 0.00 | 0 |
| 1.12 | 0.89 | -0.01 | 1 |
| 1.09 | 0.89 | -0.05 | 4 |
| 1.03 | 0.89 | -0.18 | 9 |
| 0.92 | 0.86 | -0.40 | 17 |
| 0.77 | 0.82 | -0.64 | 24 |
| 0.60 | 0.73 | -0.81 | 33 |
| 0.43 | 0.60 | -0.91 | 42 |
| 0.27 | 0.42 | -0.96 | 52 |

## 8 Bayesian hierarchical models *FM-initial* 10 – 45 for Fitters: Model comparison based on the WAIC and interpretation of marginal posterior distributions (c.f. section Results in Main Body)

**Supplementary Figure 3.**
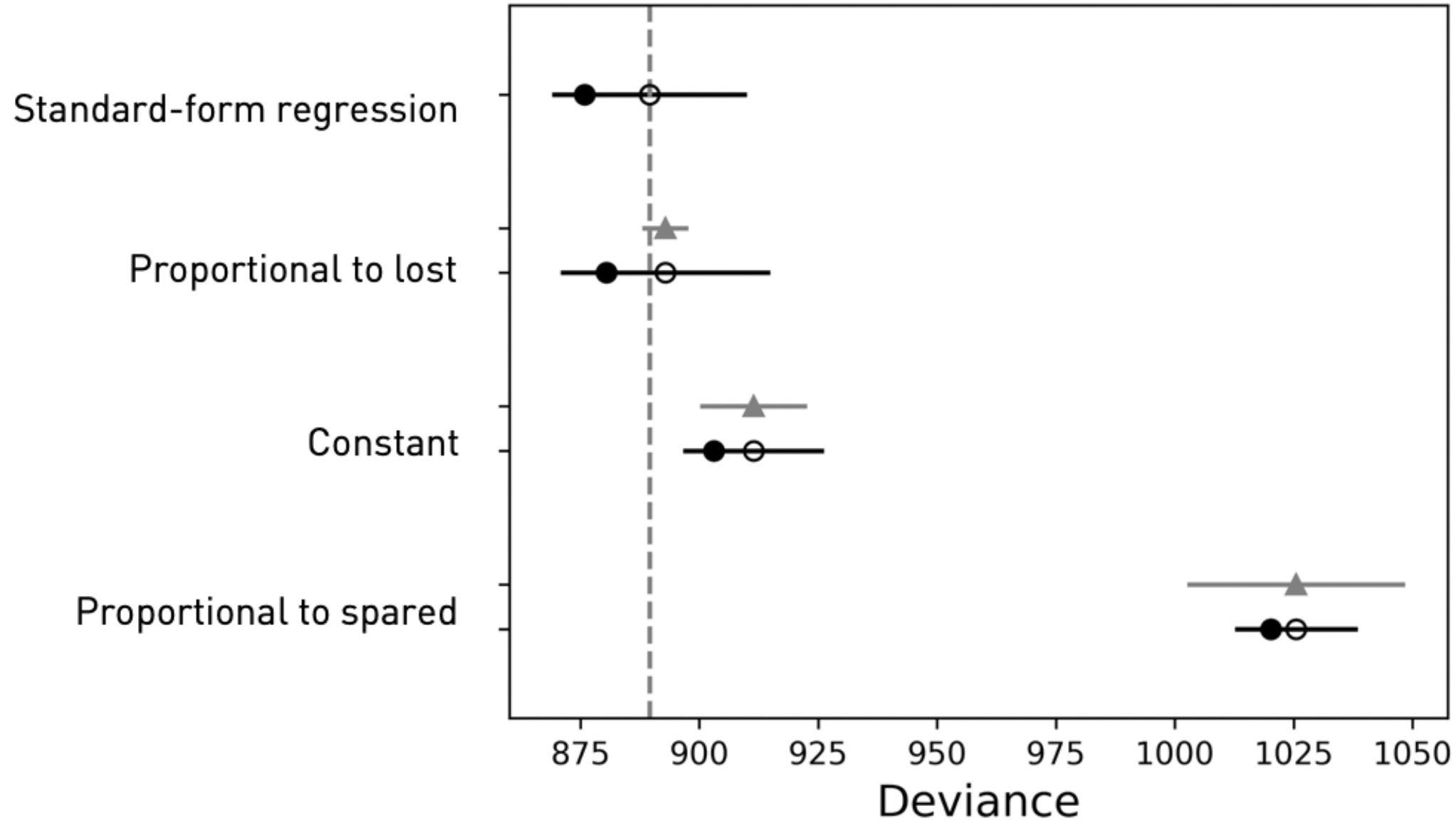
Subset *FM-Initial* 10 – 45 for *fitters*. Model comparison based on the widely applicable criterion (WAIC), resulting in a similar model ranking in comparison to the leave-one-out-cross-validations results. The standard-form regression model is top-ranked, closely followed by the proportional to lost function model and constant recovery model.

## 9 Further Discussion of Fitting Results (c.f. section Results in Main Body)

Here, we provide further interpretation of the fitting results that we obtain for our human data. There are three cases where our fitting gives parameter estimates that are just outside the range of accurate retrieval according to our ground-truth (synthetic data) simulations. This happens for change-form proportional to spared for both *fitters* alone and *fitters & non-fitters* combined, and change-form constant recovery for *fitters* alone. Importantly, these are all losing models. That is, for *fitters*, change-form proportional to spared was the worst fitting followed by change-form constant recovery, while for *fitters & non-fitters*, change-form proportional to spared was the worst model.

Since these are losing models, the key inferential error that we want to defend ourselves against is that we have *under*-estimated the R-squared and model evidence for these, since only this would alter our conclusions, as to which are the winning models. Critically, though, in all these cases, any inaccuracy in fitting would work in the opposite direction, i.e., to *over*-estimate the quality of fit. This is very simple because if the particular model was the true generating model, the correct parameter setting would be in the parameter space being fitted. The parameter values selected in the model fitting have to be the correct settings or “better” fittings than them.

Otherwise, they would not have been selected by the fitting algorithm.

Thus, in all these critical cases, the R-squared and model evidence can only be correct or *over*-estimated, *under*-estimation is not possible. However, even if there is *over*-estimation, the models still lose, confirming that the inferences we have made are sound. We discuss these three cases in a little more depth next.

### Constant Recovery

when we fit the constant recovery rule in its change formulation (see **Figure 2[C]**) to the Fitters group, we obtain an R-squared of 5.8% and a constant of 26pt (see section “Final Model Comparison (on human data) in the Subset of FM-initial 10-45”). In supplementary material section 8 (“Detailed Evaluation of Simulations”), we show synthetic data simulation results for the different models. The simulations for constant recovery, see **Supplementary Table 3f**, show that with 10-45 subsetting, we get very accurate parameter retrieval for constants up to 20 points. For a constant of 25, though, retrieval becomes less accurate and worsens further for 30 and beyond.

The central inference that we are seeking to make in our analysis of fitters is that standard-form regression has the highest R-squared and model evidence, followed by proportional to lost function and then constant recovery. Thus, as just discussed, the scenario that we wish to defend ourselves against is that we have underestimated the R-squared and model evidence for the fit of constant recovery. For the reasons just discussed, we can argue that such an underestimation has not been made. This, then, suggests that our conclusion that standard-form and proportional to lost fit better than constant recovery for the 10-45 subset of the *fitters* data, is indeed a reliable inference.

For the *Fitters & Non-fitters* combined data set, constant recovery obtains a constant of ∼20points, putting it in the range of reliable parameter retrieval.

### Proportional to Spared

When we fit the proportional to spared model in the change formulation (see **Figure 2[B]**) for both *fitters* alone and *fitters & non-fitters*, we obtain slopes that are in the upper part of the allowable range, where retrievability of parameters starts to breakdown; see **Supplementary Table 2f**, for 10-45 subsetting, which is around 60%. However, both for *fitters* and *fitters & non-fitters*, when this arises, we can use the argument just made to justify that an under-estimate of fit has not been made, supporting our inferences that models other than proportional to spared are the best fitting.

## 10 Additional subset evaluations for Fitters: Bayesian hierarchical models *FM-initial* 10 – 40 and *FM-initial* 10 – 50 (c.f. Results section of Main Body)

**Supplementary Figure 4.**
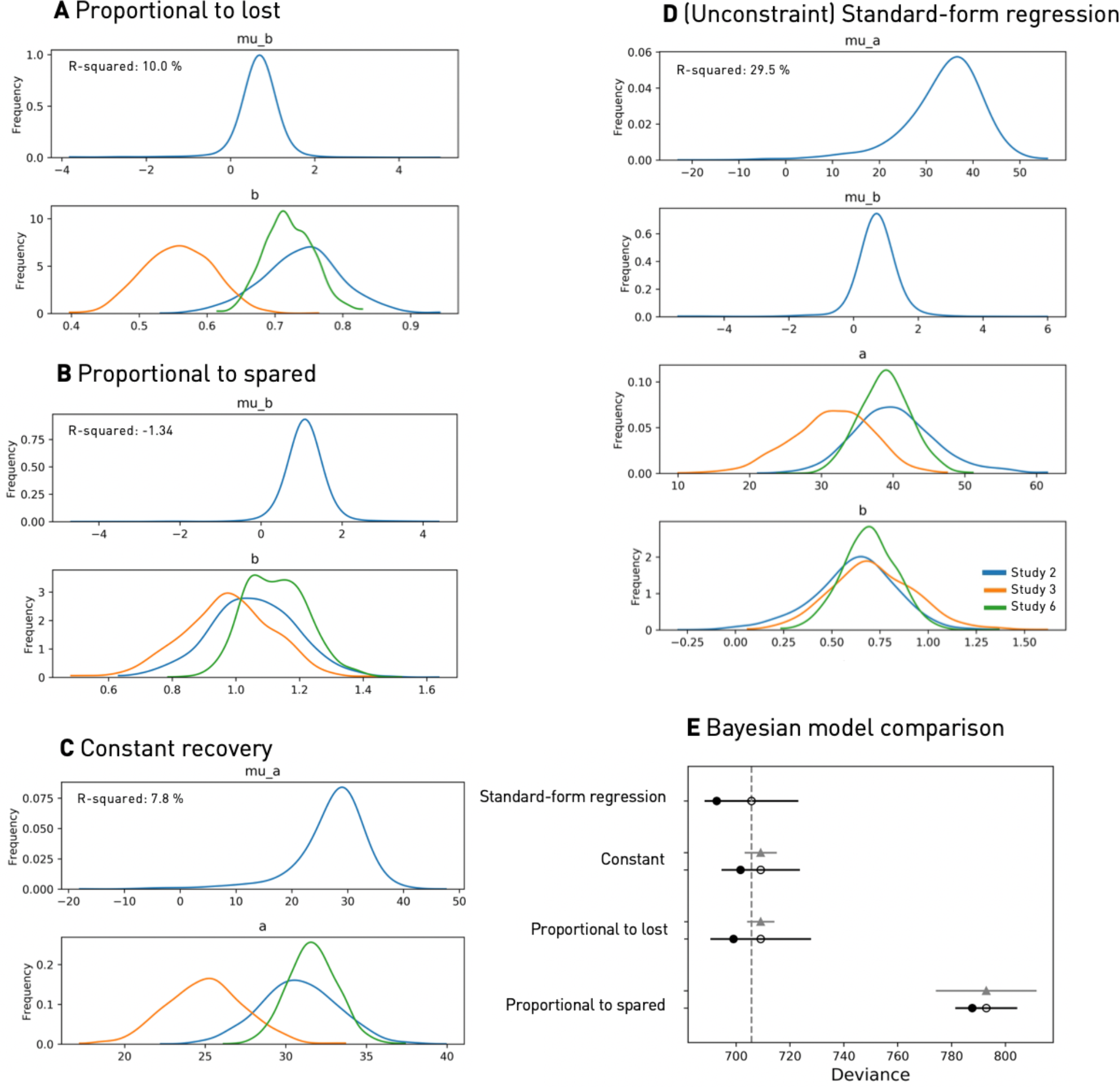
Alleviating confounds by ceiling effects and mathematical coupling: Marginal posteriors for Fitters of the hierarchical models’ parameters considering all subjects with initial FM scores between ten and 40. Final model comparison. A. Proportional to lost function recovery (formulated as in **Figure 2[A])**; B. Proportional to spared function recovery (formulated as in **Figure 2[B]**); C. Constant recovery (formulated as in **Figure 2[C]**); D. (Unconstrained) Standard-form regression; E. Final Bayesian model comparison relying on leave-one-out-cross-validation. Empty circles represent the LOOCV-Score, black error bars the corresponding standard error. The filled black circles mark the models’ in-sample deviance, grey triangles the difference to the top-ranked model, as well as the associated standard error. Lastly, the lowest LOOCV-value is indicated by the vertical dashed grey line. The standard-form regression model provides the best out-of-sample performance and is ranked first in the model comparison, closely followed by the constant and proportional to lost function recovery models.

**Supplementary Figure 5.**
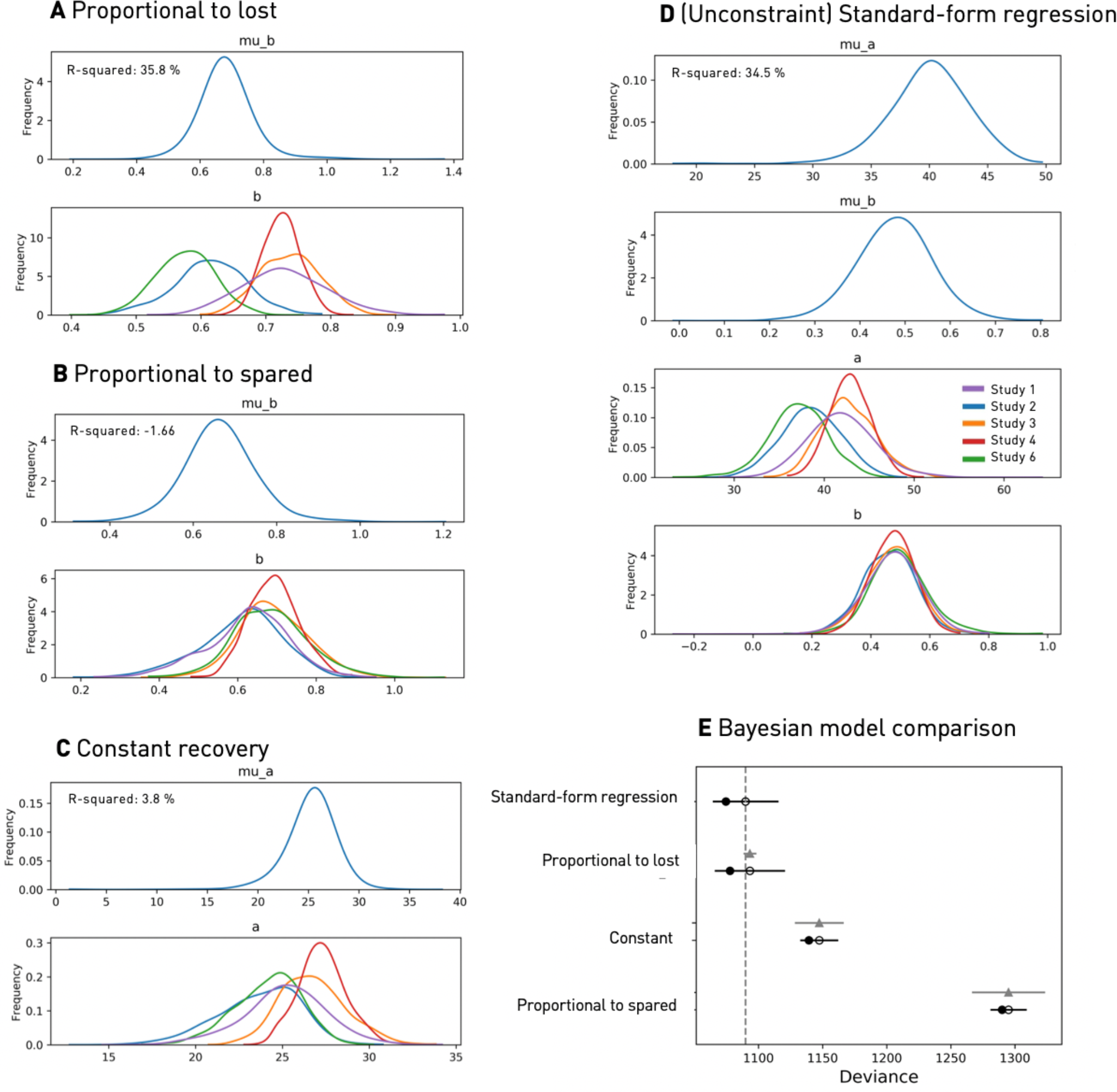
Alleviating confounds by ceiling effects and mathematical coupling: Marginal posteriors for Fitters of the hierarchical models’ parameters considering all subjects with initial FM scores between ten and 50. Final model comparison. **A**. Proportional to lost function recovery (formulated as in **Figure 2[A]**); **B**. Proportional to spared function recovery (formulated as in **Figure 2[B]**); **C**. Constant recovery (formulated as in **Figure 2[C]**); **D**. (Unconstrained) Standard-form regression; **E**. Final Bayesian model comparison relying on leave-one-out-cross-validation. Empty circles represent the LOOCV-Score, black error bars the corresponding standard error. The filled black circles mark the models’ in-sample deviance, grey triangles the difference to the top-ranked model, as well as the associated standard error. Lastly, the lowest LOOCV-value is indicated by the vertical dashed grey line. The standard-form regression model provides the best out-of-sample performance and is ranked first in the model comparison, immediately followed by the proportional to lost function recovery model. The constant recovery model follows with quite some distance, the proportional to spared recovery model is ranked last, as in the other subsets’ model comparisons.

## Footnotes

1 We use the word “function” to indicate behavioural performance, e.g. of basic motor abilities on a scale such as the Fugl Meyer.

2 For further clarification: This composite involved applying the model across the entire range of subjects, *fitters & non-fitters*. Here, we are not considering fitting of different models to different portions of the data.

